# Unveiling the hidden threat: the impact of sub-optimum treatment on acquired immunity, asymptomatic cases, and malaria dynamics

**DOI:** 10.64898/2026.03.24.26349187

**Authors:** Hemaho B. Taboe, Megan Y. Sin, Madison Pratt, Elizabeth J. Rush, Charles Mbogo, Olivia Prosper Feldman, Ruijun Zhao, Calistus N. Ngonghala

**Author notes:** Corresponding author: Calistus Ngonghala.

## Abstract

Malaria persists worldwide, exerting its greatest impact in sub-Saharan Africa. This study develops and uses a mathematical model to assess how sub-optimum versus optimum treatment of malaria drives asymptomatic infections, immunity build-up, and sustained transmission, providing insights for effective control. Fitted to case data from Kenya and Nigeria, the framework is used to quantify the burden of malaria and the additional cost associated with sub-optimum treatment. Global sensitivity analysis identifies mosquito demographic parameters, biting rates, and malaria treatment rate among major disease drivers under sub-optimum treatment, emphasizing the need for integrated strategies that improve access to optimum treatment and reduce mosquito–human contact. Model simulations show that sub-optimum treatment amplifies asymptomatic prevalence, sustaining/increasing malaria transmission and burden. Further simulations reveal that optimum treatment could avert more than one-third of infections and deaths, while asymptomatic infections contribute up to 96% (75%) of malaria-related Years Lived with Disability in Kenya (Nigeria). Cost analysis shows that optimum treatment lowers malaria burden significantly and can reduce annual total treatment costs by ≈ $12 million, underscoring the substantial economic and public health gains of limiting sub-optimum care. This study demonstrates that effective and sustained malaria control requires strengthening adherence to treatment, minimizing sub-optimum treatment, reducing mosquito–human contact, and targeting asymptomatic carriers to curb hidden transmission and reduce long-term health and economic losses.

## 1. Introduction

Malaria is a life-threatening parasitic mosquito-borne disease that has been one of the leading causes of death and illness worldwide. Spread by bites from infected female *Anopheles* mosquitoes, the disease is characterized by symptoms including febrile illness, anemia, seizures, renal failure, acute respiratory distress, and even coma [1]. Malaria can be fatal, with a mortality rate of 14.3 deaths per 100, 000 population at risk in 2022 [2]. It affects the World Health Organization (WHO) African Region disproportionately, with about 233 million (580, 000) cases (deaths) in 2022, making up 94% (95%) of the global cases (deaths) [3].

In malaria-endemic regions, repeated exposure to infection leads to the development of acquired (clinical) immunity, which suppresses parasite replication and mitigates severe symptoms, even though individuals may continue to harbor some parasites [4–10]. This immunity arises as the immune system adapts progressively to diverse parasite antigens, providing partial protection against severe disease and death [4–10]. However, this protection is not complete, and individuals can still experience mild (asymptomatic) infections, highlighting the need for continuous preventive measures [11]. Clinical immunity exists on a spectrum, with individuals experiencing severe symptoms like seizures or acute respiratory diseases at one end, and individuals with mild or asymptomatic infections at the other [4]. Generally, symptomatic individuals are more infectious than asymptomatic carriers due to higher parasitemia levels [12–15]. But, despite lower parasite loads, asymptomatic individuals can still contribute significantly to malaria transmission, especially in high-prevalence areas, by carrying parasites that remain undetected by surveillance systems for extended periods without seeking treatment [15–17]. Sub-optimum (incomplete or unconventional) treatment practices in many endemic settings, contribute to the persistence of these asymptomatic infections. Specifically, sub-optimum treatment can affect the transition from symptomatic to asymptomatic malaria significantly, as it can leave a residual parasite load that sustains subclinical infections. Moreover, sub-optimum treatment can increase the likelihood of drug resistance, further complicating malaria control efforts [18, 19].

Given these challenges, mathematical modeling studies have further elucidated how asymptomatic infection, immunity, and vector control shape malaria dynamics. For example, Cai *et al*. [20] used an SEIR framework to assess the impact of asymptomatic infection and superinfection on malaria dynamics, while Lindblade *et al*. [15] showed that asymptomatic carriage can persist for months. Koutou *et al*. [21] incorporated partial immunity and incubation delays, demonstrating delay-driven oscillations and their effects on malaria transmission. Aguilar *et al*. [22] modeled malaria systems without a recovered class and showed that asymptomatic hosts cause substantial underestimation of the basic reproduction number (*R*_0_). Keegan and Dushoff [23] analyzed clinical immunity to malaria and identified conditions for bistability, and Gebremeskel and Krogstad [24] established backward bifurcation arising from standard incidence or superinfection, highlighting the importance of combined bed-net use and treatment to malaria control. Complementary work by Ngonghala [25] used a malaria model with an asymptomatic class to demonstrate how net-efficacy decay, attrition, and insecticide resistance influence intervention success and emphasized strategies such as pyrethroid-piperonyl butoxide-enhanced nets and timely net replacement. Taboe *et al*. [26] used a malaria transmission model that incorporated heterogeneous mosquito biting and dynamic immunity to demonstrate that adult mosquito mortality is the dominant determinant of transmission intensity and that neglecting biting heterogeneity leads to inaccurate estimates of population immunity, disease burden, and intervention impact. Additional studies in [7, 8, 19, 27] capture how repeated exposure, symptomatic–asymptomatic transitions, and fluctuating immunity shape malaria prevalence and intervention effectiveness across heterogeneous immunity profiles. Unlike these modeling studies that focused on asymptomatic infection, immunity loss, superinfection, and intervention performance, other studies have focused primarily on the effects of climate change, human behavior, mosquito feeding and reproduction trends, etc., on malaria transmission and control [28–36].

The objective of this work is to develop and analyze a model framework for malaria dynamics that accounts for optimum and sub-optimum treatment of infectious individuals, asymptomatic cases, and two human immunity states (low versus high immunity) resulting from optimum versus sub-optimum treatment. The model is fitted to observed (confirmed) malaria case data from Nigeria, a region with particularly high malaria prevalence, and Kenya, a region with lower prevalence. The calibrated model is used to 1) investigate how sub-optimum treatment affects the emergence and persistence of asymptomatic infections and the role of asymptomatic cases as hidden malaria reservoirs; assess the economic burden of malaria through Disability Adjusted Life Years and the cost and economic impact of sub-optimum malaria treatment (in relation to the net gain accrued by improving treatment completion); and to explore the similarities and differences in malaria control between Kenya and Nigeria. To our knowledge, there is no model for malaria that integrates optimum and sub-optimum treatments to assess their impacts on the emergence and persistence of malaria, while also assessing the associated economic burden of malaria.

## 2. Methods

### 2.1. Model formulation

In this section, a mathematical model for malaria that accounts for low and high clinical immunity dynamics is developed. The total human population (*N*_*h*_), is divided into the following compartments: susceptible (*S*_*h*_), exposed but not yet infectious (*E*_*h*_), symptomatic infected with moderate to severe symptoms (*I*_*h*_), recovered with low immunity after optimum treatment (*R*_*w*_), re-exposed with low immunity (*E*_*w*_), moderately to severely infectious with low immunity (*I*_*wc*_), mild symptomatic to asymptomatic infectious with low immunity (*I*_*wa*_), asymptomatic infectious with high immunity after sub-optimum treatment (*I*_*sa*_), re-exposed asymptomatic with high immunity (*E*_*s*_), moderate to severe symptomatic infectious with high immunity (*I*_*sc*_), and recovered with high immunity following sub-optimum treatment (*R*_*s*_). The total vector population (*N*_*v*_) is divided into susceptible (*S*_*v*_), exposed (*E*_*v*_), and infectious (*I*_*v*_) compartments. The flow diagram of the model framework is presented in Fig. 1, while descriptions of the model variables and parameters are summarized in Tables 1-2.

**Table 1:** Brief definitions of the state variables of the model (2.1).

| State variable | Description of state variable |
| --- | --- |
| $S_h (S_v)$ | Susceptible humans (mosquitoes), who are at risk of infection but not infected yet. |
| $E_h (E_v)$ | Exposed humans (mosquitoes), who are infected but not infectious yet. |
| $I_h (I_v)$ | Infectious humans with moderate to severe symptoms (infectious mosquitoes). |
| $R_w (R_s)$ | Recovered humans with low (high) immunity due to optimum (sub-optimum) treatment. |
| $E_w (E_s)$ | Re-exposed humans with low (high) immunity. |
| $I_{wc} (I_{sc})$ | Moderate to severe symptomatic infectious humans with low (strong) immunity. |
| $I_{wa} (I_{sa})$ | Asymptomatic infectious humans with zero to mild symptoms and low (strong) immunity. |

**Table 2:** Descriptions of parameters. The subscript *j* = *w* (*j* = *s*) denotes low (high) immunity, *k* = *c* (*k* = *a*) denotes symptomatic (asymptomatic), while *H, M*, and yr denote human, mosquito, and year, respectively.

| Parameter | Description of parameter | Unit |
| --- | --- | --- |
| $\Lambda_h (\Lambda_v)$ | Human (mosquito) recruitment rate. | $H \text{ yr}^{-1} (M \text{ yr}^{-1})$ |
| $1/\mu_h (1/\mu_v)$ | Average human (mosquito) lifespan. | $\text{yr}^{-1}$ |
| $\delta_h (\delta_w) (\delta_{sc})$ | Malaria-induced death rate for the $I_h$ (low) (high) immunity human groups. | $\text{yr}^{-1}$ |
| $1/\sigma_h (1/\sigma_v)$ | Intrinsic (extrinsic) incubation period of malaria. | $\text{yr}^{-1}$ |
| $1/\sigma_{wa} (1/\sigma_{sa})$ | Incubation period of asymptomatic individuals with low (high) immunity. | $\text{yr}^{-1}$ |
| $1/\sigma_{wc} (1/\sigma_{sc})$ | Incubation period of symptomatic individuals with low (high) immunity. | $\text{yr}^{-1}$ |
| $1/\rho_w (1/\rho_s)$ | Average duration of immunity for $R_w (R_s)$ humans. | $\text{yr}^{-1}$ |
| $1/\gamma_{sa}$ | Average duration of malaria for individuals in the $I_{sa}$ class. | $\text{yr}^{-1}$ |
| $1/\gamma_{wa}$ | Average duration of malaria for individuals in the $I_{wa}$ class. | $\text{yr}^{-1}$ |
| $\gamma_{jc} (\phi_{jc})$ | Optimum (sub-optimum) treatment rate of individuals in the $I_{jc}$ class. | $\text{yr}^{-1}$ |
| $\psi_{wa} (\psi_{sa})$ | Rate at which $I_{wa} (I_{sa})$ asymptomatic humans become symptomatic. | $\text{yr}^{-1}$ |
| $\xi_{sa}$ | Waning rate of immunity in the strong immunity group. | $\text{yr}^{-1}$ |
| $q_w (q_s)$ | Infectious fraction with low (high) immunity who become asymptomatic. | Dimensionless |
| $1/\phi_{h1}$ | Average duration of malaria treatment. | $\text{yr}^{-1}$ |
| $1/\phi_{h2}$ | Average time from symptomatic stage to asymptomatic if not treated. | $\text{yr}^{-1}$ |
| $p(\varepsilon)$ | Proportion of individuals who seek (complete optimum) treatment. | Dimensionless |
| $\tau$ | Efficacy of malaria treatment. | Dimensionless |
| $\beta_{vh} (\beta_{hv})$ | Average number of bites that a mosquito places on susceptible (infectious) humans per unit time. | $\text{yr}^{-1}$ |
| $p_{hv}$ | Transmission probability from infectious humans to mosquitoes. | Dimensionless |
| $p_{vh}$ | Transmission probability from infectious mosquitoes to humans. | Dimensionless |
| $\theta$ | Modification factor of asymptomatic transmission. | Dimensionless |

**Figure 1.**
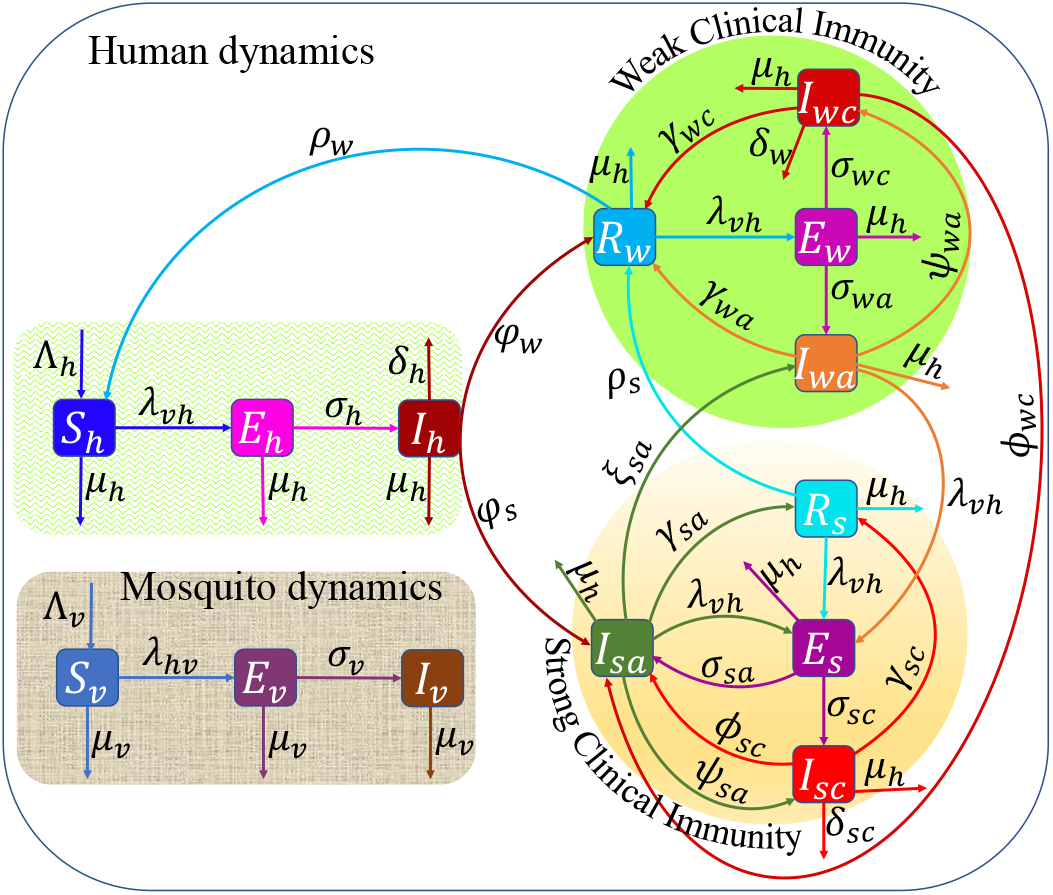
Schematics of the malaria model. Conceptual framework of the model (2.1) depicting the flow of humans and mosquitoes between different compartments representing disease status. The flow rate *φ*_*s*_ = *pϕ*_*h*1_(1 − *ετ* ) + (1 − *p*)*ϕ*_*h*2_ and *φ*_*w*_ = *ετpϕ*_*h*1_, where 1/*ϕ*_*h*1_ is the average duration of treatment and 1/*ϕ*_*h*2_ is the average time for symptomatic infectious individuals to become asymptomatic if they do not receive treatment. It is assumed that *γ*_*wc*_ = *γ*_*sc*_ = *φ*_*w*_, *ϕ*_*wc*_ = *ϕ*_*sc*_ = *φ*_*s*_, *σ*_*wa*_ = *q*_*w*_*σ*_*w*_ = *q*_*w*_*σ*_*h*_, *σ*_*sa*_ = *q*_*s*_*σ*_*s*_ = *q*_*s*_*σ*_*h*_, *σ*_*wc*_ = (1 − *q*_*w*_)*σ*_*s*_ = (1 − *q*_*w*_)*σ*_*h*_, and *σ*_*sc*_ = (1 − *q*_*s*_)*σ*_*s*_ with *q*_*s*_ > *q*_*w*_ since individuals with high immunity are more likely to become asymptomatic than those with low immunity. Descriptions of the state variables and parameters are given in the text and summarized in Tables 1 and 2, respectively.

The model assumes no vertical transmission of malaria, so all human (mosquito) births flow into the susceptible class at rate Λ_*h*_ (Λ_*v*_). Humans (mosquitoes) in each of the compartments die naturally at per capita rate *µ*_*h*_ (*µ*_*v*_). Susceptible humans (mosquitoes) become exposed at a rate *λ*_*vh*_ = *β*_*vh*_*p*_*vh*_*I*_*v*_/*N*_*h*_ (*λ*_*hv*_ = *β*_*hv*_*p*_*hv*_[*I*_*h*_+*I*_*wc*_+*θ*(*I*_*wa*_+ *I*_*sa*_) + *I*_*sc*_]/*N*_*h*_), where *β*_*vh*_ (*β*_*hv*_) is the average number of mosquito bites on a susceptible (infected) human per time, *p*_*vh*_ is the probability that an infectious mosquito infects a susceptible human, and *p*_*hv*_ is the probability that an infectious human infects a susceptible mosquito. The modification parameter 0 < *θ* < 1 accounts for the reduced infectivity of asymptomatic infectious humans, primarily due to the lower levels of parasitaemia observed in these individuals. Exposed mosquitoes become infectious at rate, *σ*_*v*_. Initially infected humans become symptomatically infectious at per capita rate, *σ*_*h*_, while symptomatically infectious humans recover with weak immunity after optimum treatment at per capita rate, *ψ*_*w*_ = *ετpϕ*_*h*1_. Here, *p* is the fraction of symptomatic infectious humans receiving treatment, *ε* the fraction who complete it, *τ* the fraction of effective treatments, and *ϕ*_*h*1_ is the reciprocal of the average duration of malaria treatment. Symptomatic infectious individuals who receive incomplete (suboptimum) treatment enter the asymptomatic infectious class at per capita rate *ψ*_*s*_ = *pϕ*_*h*1_(1 − *ετ* ) + (1 − *p*)*ψ*_*h*2_, where 1/*ψ*_*h*2_ is the average time for symptomatic infectious individuals to become asymptomatic if they receive unconventional or no treatment. Individuals who recover with low/weak immunity through optimum treatment lose their immunity at rate *ρ*_*w*_ to join the susceptible class, or are reinfected at a rate *λ*_*vh*_ to enter the low immunity exposed class. Members of the low immunity exposed class either become asymptomatic (*I*_*wa*_) at rate *σ*_*wa*_, or symptomatic infectious (*I*_*wc*_) at rate *σ*_*wc*_. Low immunity symptomatically infectious individuals die from malaria at rate *δ*_*w*_, recover from infection with low immunity after receiving optimum treatment at rate *γ*_*w*_, or transition to the high immunity asymptomatic infectious class *I*_*sa*_ at per capita rate (*ϕ*_*wc*_), if they receive sub-optimum treatment. Notice that we assume the optimum treatment rate remains the same for any individual receiving treatment. Low immunity asymptomatic infectious individuals (*I*_*wa*_) are reinfected at a rate *λ*_*vh*_ to enter the high immunity exposed class, recover with low immunity at rate (*γ*_*wa*_) to join the *R*_*w*_ class or at rate *ψ*_*wa*_, to join the low immunity symptomatic infectious class (*I*_*wc*_). Individuals in the high-immunity asymptomatic infectious class (*I*_*sa*_) join the high immunity symptomatic infectious state (*I*_*sc*_) at rate *ψ*_*sa*_, recover into the high immunity class (*R*_*s*_) at rate *γ*_*sa*_, join the low immunity class (*I*_*wa*_) when their immunity wanes at rate, *ξ*_*sa*_, or become reinfected at rate *λ*_*vh*_ to join the high immunity exposed class *E*_*s*_. Individuals in the high-immunity recovery class transition to the low-immunity recovered class (*R*_*w*_) as their immunity wanes at rate *ρ*_*s*_, or they become reinfected at rate *λ*_*vh*_ and move into the high-immunity exposed class. High-immunity exposed humans develop symptoms at rate *σ*_*sc*_ to join the *I*_*sc*_ class, or become asymptomatic at rate *σ*_*sa*_ to rejoin the *I*_*sa*_ class. Symptomatic individuals in this high immunity group die due to disease at rate *δ*_*sc*_, progress to the high immunity recovery class (*R*_*w*_) at rate *γ*_*sc*_, upon optimum treatment, or return to the asymptomatic class (*I*_*sa*_) at rate *ϕ*_*sc*_, after receiving sub-optimum treatment.

### 2.2. Model equations

Using the variables (Table 1), parameters (Table 2), and the flow diagram (Fig. 1), we derive the model system:

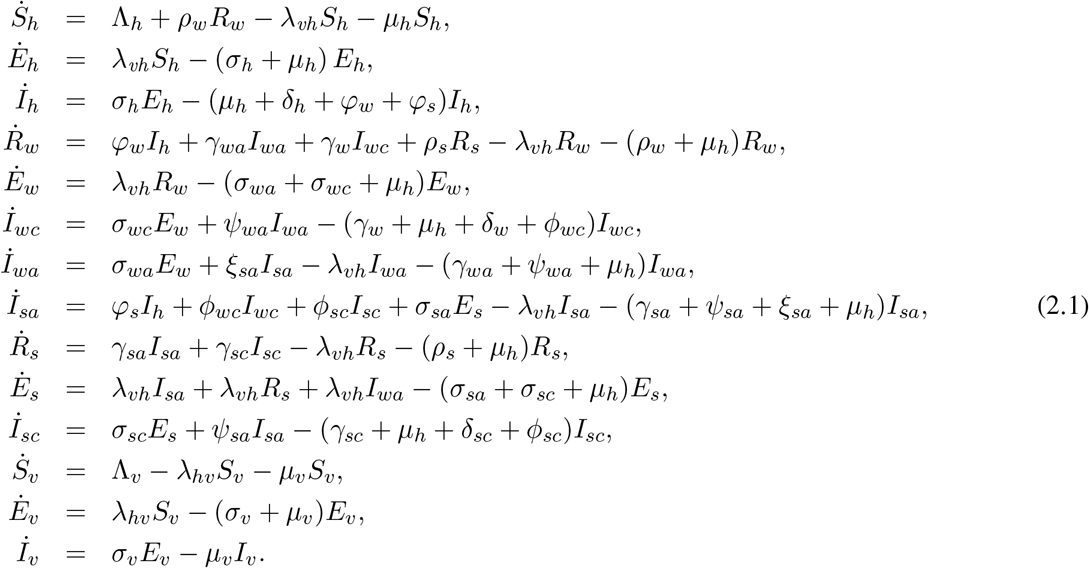

Let *X* = (*S*_*h*_, *E*_*h*_, *I*_*h*_, *R*_*w*_, *E*_*w*_, *I*_*wc*_, *I*_*wa*_, *I*_*sa*_, *R*_*s*_, *E*_*s*_, *I*_*sc*_, *S*_*v*_, *E*_*v*_, *I*_*v*_). Then the model (2.1) will be studied together with the initial condition *X*(0) = (*S*_*h*0_, *E*_*h*0_, *I*_*h*0_, *R*_*w*0_, *E*_*w*0_, *I*_*wc*0_, *I*_*wa*0_, *I*_*sa*0_, *R*_*s*0_, *E*_*s*0_, *I*_*sc*0_, *S*_*v*0_, *E*_*v*0_, *I*_*v*0_), where all initial values are non-negative. Consequently, all solutions of system (2.1) remain non-negative for all non-negative time. In the rest of the paper, we use the following parameter groupings for notational convenience:

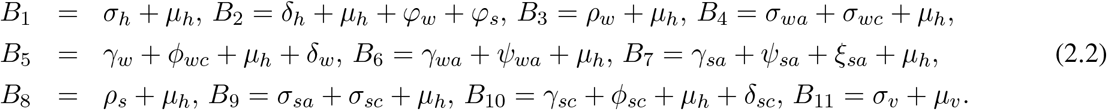

### 2.3. Well-Posedness of the model

The dynamics of the total human and mosquito populations are governed by the equations: 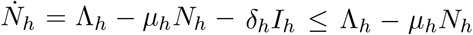 and 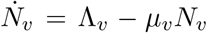, respectively. Thus, the growth of *N*_*h*_ and *N*_*v*_ are characterized by 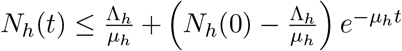, and 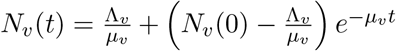, where *N*_*h*_(0) and *N*_*v*_(0) are the respective initial total human and mosquito populations. Observe that 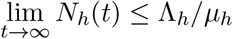 and 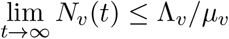. Hence, 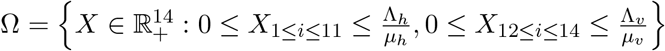 is a positively-invariant region (i.e., solutions that start in Ω remain in Ω, for all positve time) and attracts all solutions of the model. Analysis of the model will be performed within this epidemiological and mathematically feasible region.

## 3. Results

### 3.1. Analytical results

#### 3.1.1. Disease free equilibrium and basic reproduction number

The disease-free equilibrium (*DFE*) of the model (2.1) defined as the steady state of the model in which no infection is present in the population (i.e., when all infected compartments are zero) is given by 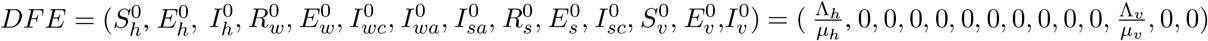. This equilibrium will be used together with the next generation matrix approach to compute the reproduction number (ℛ_0_) of the model, defined as the expected number of new human (mosquito) infections generated by a single infectious case (human or mosquito) introduced into a fully susceptible human or mosquito population, accounting for the full transmission cycle between humans and mosquitoes. It reflects the combined effects of the mosquito biting rate, mosquito survival, infection probabilities, and parasite development, etc., and serves as a threshold indicator determining whether malaria will die out. Using the Next Generation Operator technique, the reproduction number is given by the spectral radius of the matrix *FV* ^−1^ (i.e., ℛ_0_ = *ρ*(*FV* ^−1^)), where *F* is the new infection matrix and *V* is the transition matrix. For the model described by Eqs. (2.1),

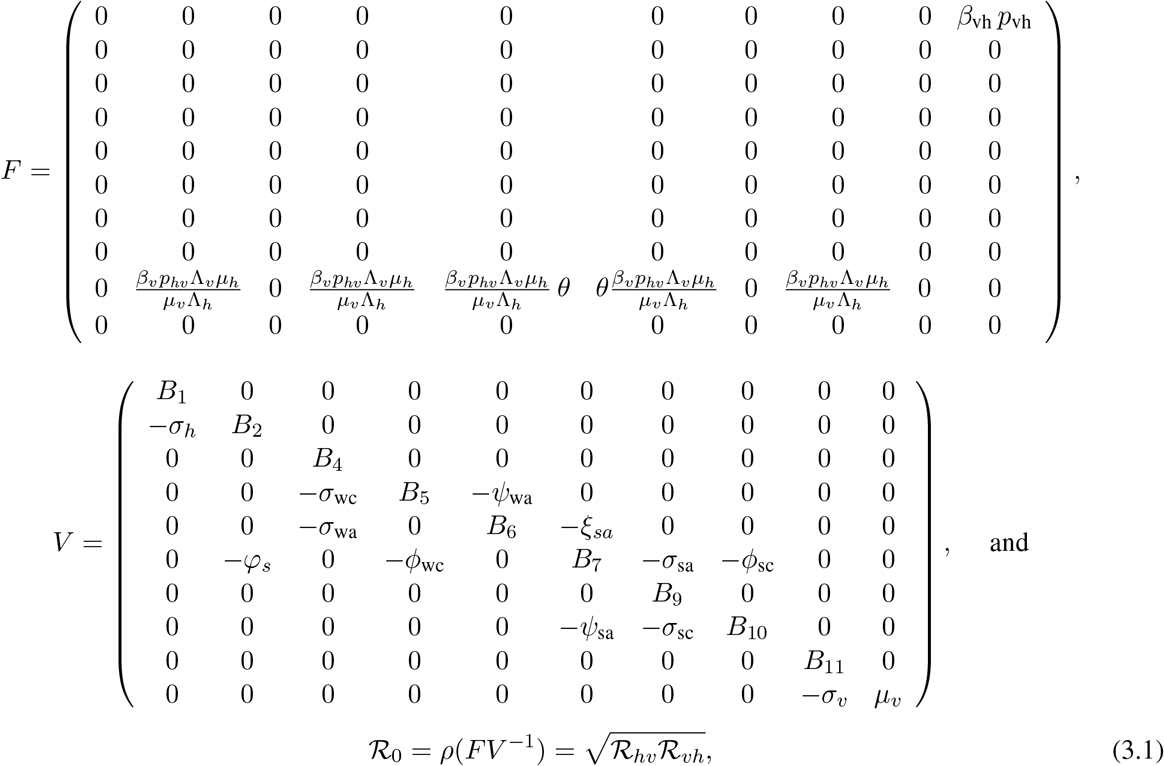

where 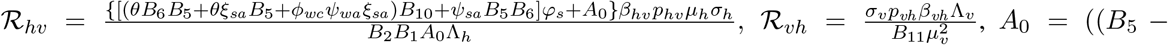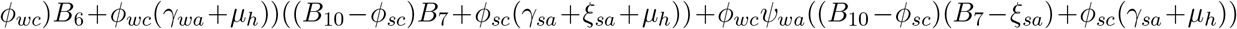, and the *B*_*i*_’s, *i* = 1, 2, 3, …, 11 are defined in Eq. (2.2). It should be noted from Eq. (2.2) that *B*_5_ − *ϕ*_*wc*_, *B*_7_ − *ξ*_*sa*_, and *B*_10_ − *ϕ*_*sc*_ are positive. Also, it should be noted that the expression for ℛ_0_ has been written in a form that highlights the human-to-mosquito transmission pathway (ℛ_*hv*_) and the mosquito-to-human transmission route (ℛ_*vh*_).

### 3.2. Model parametrization

Some parameters of the model (2.1) are known or can be calculated from existing demographic and epidemiological data. However, other parameters are either unknown or not directly observable. The unknown parameters will be estimated by fitting the model to data from Kenya and Nigeria, ensuring plausible and reliable projections.

#### 3.2.1. Fixed or Known Parameters

The parameters *µ*_*v*_, *ε, τ, q*_*s*_, *q*_*w*_, *σ*_*v*_, and *µ*_*h*_ are drawn from previous empirical studies, Λ_*h*_ is estimated from population data, while Λ_*v*_, *ϕ*_*h*1_, *ϕ*_*h*2_, *σ*_*h*_, *σ*_*wa*_, *σ*_*sa*_, *σ*_*wc*_, and *σ*_*sc*_ are are calculated from known information. Specifically, the average lifespan of Kenyans (Nigerians) is 65 (53) *years*. Therefore, we set *µ*_*h*_ for Kenyans (Nigerians) to 1/65 ≈ 0.0154 (1/53 ≈ 0.01887) *per year*. Since the human incubation period is ≈ 18.5 *days* [37], *σ*_*h*_ = (1/18.5)365 ≈ 19.72973. We assume that the incubation period is the same irrespective of the immunity level, i.e., *σ*_*h*_ = *σ*_*s*_ = *σ*_*w*_. The recruitment rate (Λ_*h*_) for both countries is obtained by fitting the net population to the corresponding population data from [38]. For Kenya, the best functional form that describes the growth of the population is 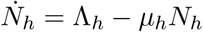 with Λ_*h*_ = 1, 759, 200 *persons per year* obtained from the fitting (Fig. 1(a) of the supplementary information (SI)). Nigeria’s population grows exponentially (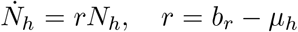, where *b*_*r*_ is the average crude birth rate. Fitting yields *b*_*r*_ = 0.0437 *per year* (Fig. 1(b) of the SI), giving a recruitment rate Λ_*h*_ = 4,672,463 *persons per year*. A data-driven estimate of human recruitment is more credible than arbitrary or equilibrium assumption approaches that can underestimate growth and mispredict disease incidence (particularly with asymptomatic cases), especially for a known growing population like Nigeria. For Λ_*v*_, we set the ratio of the mosquito birth rate to human birth rate to be 8 : 1 based on findings from [39]. Since the average lifespan of a mosquito is 14 days [37, 40–42] we set 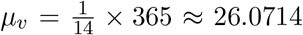 *per year*. Malaria treatment takes about 2 weeks. Therefore, *ϕ*_*h*1_ = 365 × 1/14 ≈ 26.0714 *per year*. With *q*_*s*_ = 0.893 > *q*_*w*_ = 0.648 as in [43], we set *σ*_*wa*_ = *q*_*w*_*σ*_*w*_ = *q*_*w*_*σ*_*h*_ = 0.648 × (365/18.5) ≈ 12.78487, *σ*_*sa*_ = *q*_*s*_*σ*_*s*_ = *q*_*s*_*σ*_*h*_ = 0.893 × (365/18.5) ≈ 17.61865, *σ*_*wc*_ = (1 − *q*_*w*_)*σ*_*s*_ = (1 − *q*_*w*_)*σ*_*h*_ = (1 − 0.648) × (365/18.5) ≈ 6.94486, and *σ*_*sc*_ = (1 − *q*_*s*_)*σ*_*s*_ = (1 − 0.893) × (365/18.5) ≈ 2.11108. Studies in [44] show that symptomatic individuals who do not receive treatment within two months become asymptomatic. Based on this, we set *ϕ*_*h*2_ = 365 × 1/60 *per year*. It is assumed that the disease-induced death rate is the same for all the symptomatic infectious individuals (*I*_*h*_, *I*_*wc*_, and *I*_*sc*_), i.e., *δ*_*h*_ = *δ*_*wc*_ = *δ*_*sc*_. Also, it is assumed that mosquitoes bite infectious humans at twice the rate they bite susceptible humans, supported by empirical studies showing strong preference for infected individuals [45, 46]. The values of the fixed parameters for Kenya and Nigeria and their sources are summarized in Table 3.

**Table 3:** Fixed parameter values for Kenya and Nigeria used in the analysis of model (2.1), along with their descriptions, symbols, units, and sources. The subscript *j* ∈ {*h, s, w*}.

| Parameter (description, symbol, and unit) | Kenya | Nigeria | Reference |
| --- | --- | --- | --- |
| Human recruitment rate ($\Lambda_h$ humans *per year*) | 1,759,200 | 4,672,463 | Estimated |
| Human natural mortality rate ($\mu_h$ *per year*) | 0.0154 | 0.01887 | [37, 40–42] |
| Mosquito recruitment rate ($\Lambda_v$ mosquitoes *per year*) | 14,073,600 | 37,379,704 | Calculated |
| Mosquito natural mortality rate ($\mu_v$ *per year*) | 26.07143 | 26.07143 | [37, 40–42] |
| Reciprocal of extrinsic incubation period ($\sigma_v$ *per year*) | 31.75500 | 31.75500 | [47] |
| Reciprocal of intrinsic incubation period ($\sigma_j$ *per year*) | 19.72973 | 19.72973 | [37] |
| Reciprocal of intrinsic incubation period ($\sigma_{wa}$ *per year*) | 12.78487 | 12.78487 | Calculated |
| Reciprocal of intrinsic incubation period ($\sigma_{sa}$ *per year*) | 17.61865 | 17.61865 | Calculated |
| Reciprocal of intrinsic incubation period ($\sigma_{wc}$ *per year*) | 6.94486 | 6.94486 | Calculated |
| Reciprocal of intrinsic incubation period ($\sigma_{sc}$ *per year*) | 2.11108 | 2.11108 | Calculated |
| Malaria-induced death rate ($\delta_h, \delta_{wc}, \delta_{sc}$ *per year*) | 0.03290 | 0.03290 | [47] |
| Treatment efficacy ($\tau$) | 0.90000 | 0.90000 | [2] |
| Proportion completing treatment ($\varepsilon$) | 0.65000 | 0.65000 | [48] |
| Immunity waning rate ($\rho_w, \rho_s$ *per year*) | 4.05556 | 4.05556 | [37, 40–42] |
| Low-immunity infectious fraction becoming asymptomatic ($q_w$) | 0.64800 | 0.64800 | [43] |
| High-immunity infectious fraction becoming asymptomatic ($q_s$) | 0.89300 | 0.89300 | [43] |
| Reciprocal of average treatment duration ($\phi_{h1}$ *per year*) | 26.0714 | 26.0714 | [49] |
| Reciprocal of average symptom duration ($\phi_{h2}$ *per year*) | 6.0833 | 6.0833 | [44] |

#### 3.2.2. Model fitting and estimation of unknown parameters

The parameters *ψ*_*wa*_, *ψ*_*sa*_, *p*_*hv*_, *p*_*vh*_, *β*_*vh*_, *β*_*hv*_, *γ*_*wa*_, *γ*_*sc*_,, *γ*_*sa*_, *γ*_*wc*_, *ϕ*_*sc*_, *ϕ*_*wc*_, *θ*, and *p* are estimated by fitting the model (2.1) to confirmed annual malaria case data sets for Kenya and Nigeria from 2002–2022 [50], using a nonlinear least squares method. This is achieved by minimizing the sum of the squared differences between the predicted annual cases from the model and the actual annual case data for the two countries. The minimization is implemented through the *lsqcurvefit* function in MATLAB R2023b. Goodness of fit is assessed through the residual sum of squares, which quantifies how well the model’s predictions align with the observed data and also measures the overall discrepancy between predicted and actual values. The estimated parameter values are presented in Table 4. Figure 2 shows the model fit using Kenyan data (Fig. 2(a)) and Nigerian data (Fig. 2(c)), while simulations using the estimated parameters show an excellent match between the model output and observed cumulative malaria cases (Fig. 2(b,d)). Residual dynamics for Kenya and Nigeria are shown in Fig. 2(c) and (f), respectively. The random residual patterns indicate good model fit to the data, which was further confirmed using MATLAB’s *runstest*.

**Table 4:** Descriptions of parameters, along with their symbols, units, and estimated values for Kenya and Nigeria.

| **Parameter (description, symbol, and unit)** | **Value (Kenya)** | **Value (Nigeria)** |
| --- | --- | --- |
| Transition rate from $I_{wa}$ to $I_{wc}$ ($\psi_{wa}$ per year) | 0.05005 | 0.05008 |
| Transition rate from $I_{sa}$ to $I_{sc}$ ($\psi_{sa}$ per year) | 309.99755 | 309.87890 |
| Human-to-mosquito transmission probability ($p_{hv}$) | 0.01267 | 0.00742 |
| Mosquito-to-human transmission probability ($p_{vh}$) | 0.74044 | 0.83700 |
| Mosquito biting rate of noninfectious humans ($\beta_{vh}$ per year) | 509.90840 | 868.84844 |
| Mosquito biting rate of infectious humans ($\beta_{hv}$ per year) | 1019.81680 | 1702.30986 |
| Optimum treatment-induced recovery rate ($\gamma_{wc}, \gamma_{sc}, \psi_w$ per year) | 2.30165 | 1.69295 |
| Recovery rate of weakly immune asymptomatic humans ($\gamma_{wa}$ per year) | 0.04119 | 0.05000 |
| Recovery rate of strongly immune asymptomatic humans ($\gamma_{sa}$ per year) | 0.04487 | 0.15604 |
| Proportion of individuals who seek treatment ($p$) | 0.15091 | 0.11100 |
| Sub-optimum treatment rate of $I_{sc}$ and $I_{wc}$ humans ($\phi_{sc}, \phi_{wc}$ per year) | 6.79809 | 6.60906 |
| Waning rate of immunity in strong immunity group ($\xi_{sa}$ per year) | 55.45611 | 11.40523 |
| Modification factor of asymptomatic transmission ($\theta$) | 0.99998 | 0.99988 |
| Reproduction number ($\mathcal{R}_0$) | 1.33723 | 1.34120 |

**Figure 2.**
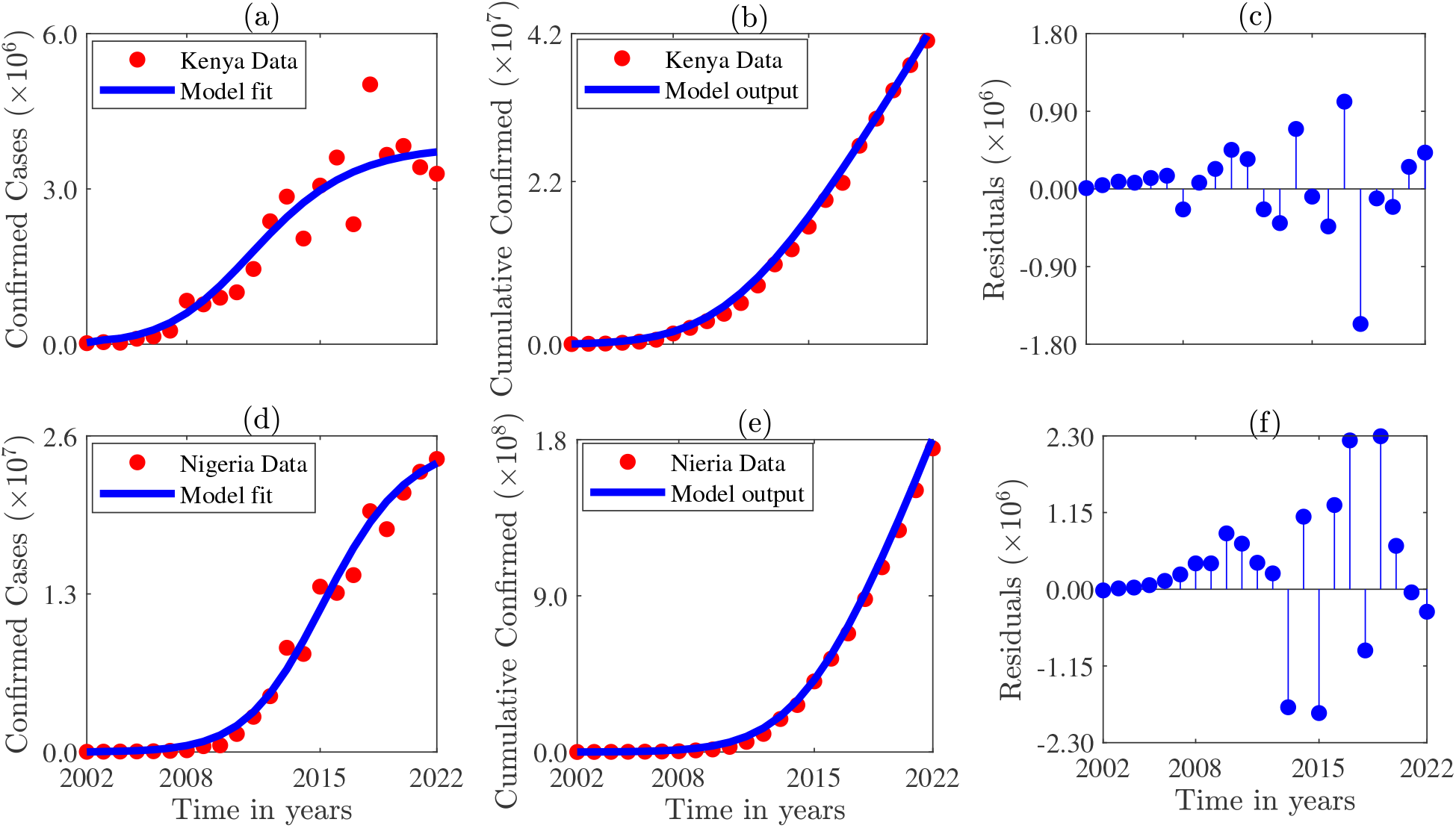
Illustration of model fitting using yearly data from (a) Kenya and (d) Nigeria, along with validation plots comparing model-predicted cumulative confirmed cases generated using the fixed and estimated parameters (Tables 3-4), with cumulative reported data for (b) Kenya and (e) Nigeria. The residuals (i.e., the difference between the model prediction and the observed data) are depicted by (c) and (f) for Kenya and Nigeria respectively. The initial conditions are presented in Table 1 of the SI.

Our fitting shows a lower human-to-mosquito than mosquito-to-human transmission probability (i.e., *p*_*hv*_ < *p*_*vh*_), which is consistent with [51] and the near-100% efficiency of *Anopheles* mosquitoes in transmitting malaria parasites to humans during blood feeding. The estimated recovery rates indicate that symptomatic humans recover faster than asymptomatic individuals, implying longer infectious periods for asymptomatic cases. Additionally, optimally treated individuals recover faster than sub-optimally treated individuals (i.e., *γ*_*wa*_ < *γ*_*sa*_). Furthermore, the estimated reproduction number for Nigeria (ℛ_0_ = 1.341) is only slightly higher than that for Kenya (ℛ_0_ = 1.337), indicating that the high burden of malaria in Nigeria might be related to its large population size.

### 3.3. Numerical results

#### 3.3.1. Sensitivity and uncertainty analysis

Global sensitivity analysis (GSA) identifies influential parameters driving variability and uncertainty in complex model outputs, thereby supporting model robustness, interpretation, and prioritization of key factors for intervention. Among GSA methods, the extended Fourier Amplitude Sensitivity Test (eFAST) is preferred to the Latin Hypercube Sampling–Partial Rank Correlation Coefficient (LHS–PRCC) approach because it quantifies both first-order effects and higher-order interactions, providing a more complete characterization of parameter influence [52–54]. Moreover, eFAST efficiently captures nonlinear and non-monotonic relationships common in biological systems, whereas LHS–PRCC is best suited to linear or monotonic settings, making eFAST a more comprehensive and reliable framework for sensitivity assessment and decision support.

Using the fixed and estimated parameter values in Tables 3–4, eFAST is applied to perform a global sensitivity analysis of the model (2.1). The analysis evaluates how parameters influence three key outcomes: exposed humans, exposed mosquitoes, and asymptomatic infectious humans due to sub-optimum treatment. These outcomes are assessed at time, *t* = 40 years. The analysis uses 5 search curves, 35 parameters (including one dummy), and 65 samples per curve, yielding a total of 11, 375 simulations. For Kenya, first-order sensitivity shows that the exposed human population (*E*_*h*_) is primarily influenced by mosquito natural mortality (*µ*_*v*_), mosquito biting rates (*β*_*vh*_, *β*_*hv*_), transmission probabilities (*p*_*vh*_, *p*_*hv*_), the vector incubation rate (*σ*_*v*_), the sub-optimum treatment rates (*ϕ*_*sc*_, *ϕ*_*wc*_), the waning rate of strong immunity (*ξ*_*sa*_), and the transition rate from asymptomatic to symptomatic infection (*ψ*_*sa*_) (Fig. 3(a)). The strongest driver is the probability of transmitting the pathogen from mosquitoes to humans, followed by the mosquito death rate and the probability of transmission from humans to mosquitoes. These findings highlight the importance of interventions that increase vector mortality such as insecticide-treated nets (ITNs) and indoor residual spraying (IRS). Also, the results highlight the critical role of treatment; timely diagnosis and effective treatment reduce the infectious reservoir and transmission, underscoring the need for improved access to diagnostics and optimum treatment for long-term malaria control. Furthermore, mosquito biting rates are influential during peak transmission and at equilibrium, reinforcing the role of measures such as ITNs, larval control, and environmental management to reduce mosquito breeding sites and the frequency of mosquito bites. Total-order sensitivity of *E*_*h*_ reveals no additional influential parameters beyond the first-order effects, with total effects exceeding first-order values (Fig. 3(a,d)). Total order sensitivity of *E*_*h*_ show no additional parameters to those exhibiting significant first order sensitivity effects (Fig. 3(d)), with overall total effects exceeding first order effects (comparing Fig. 3 (a) and (d)).

**Figure 3.**
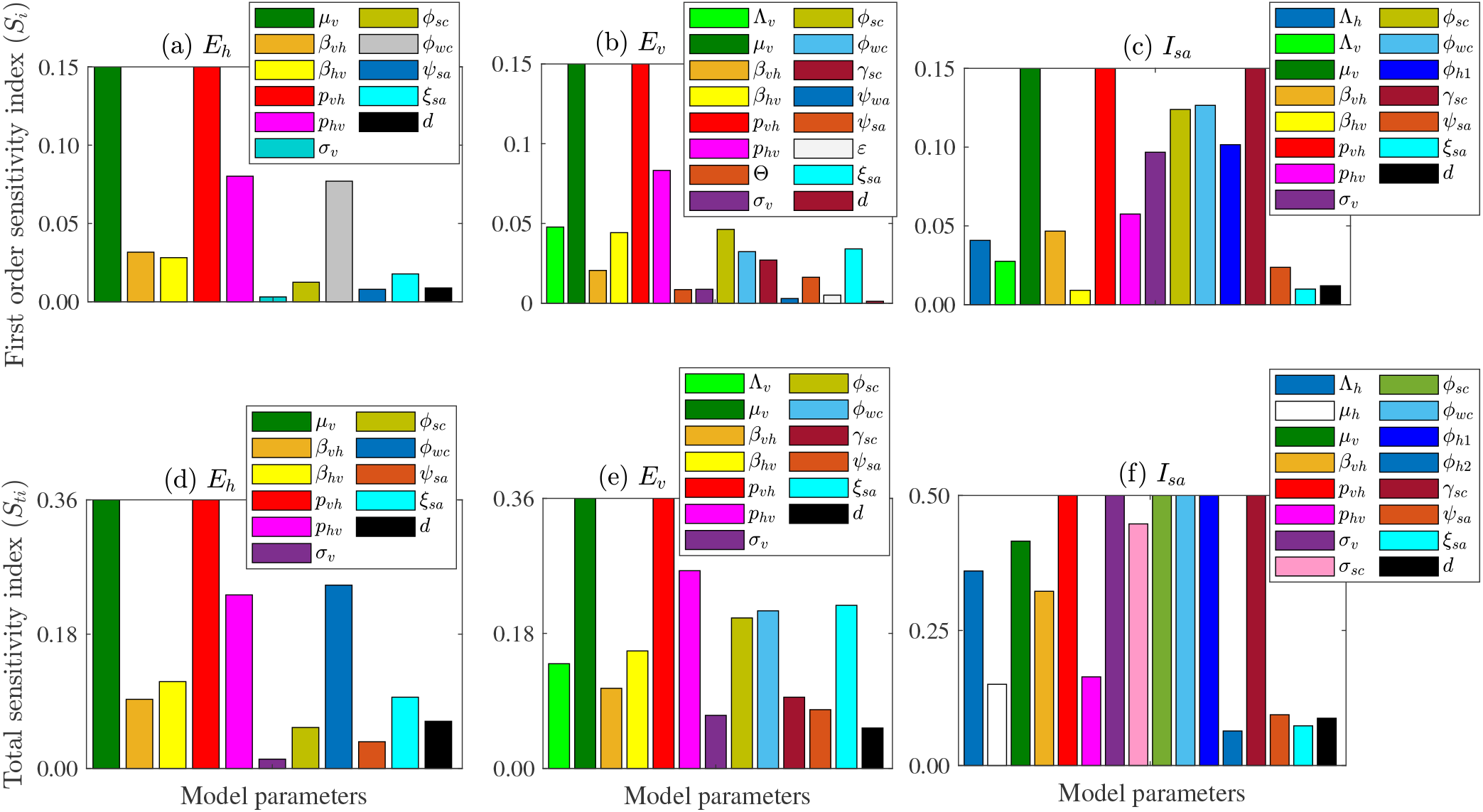
Global sensitivity analysis of the model (2.1) using the extended Fourier Amplitude Sensitivity Test (eFAST) approach. First order sensitivity of the (a) exposed human population (*E*_*h*_), (b) exposed mosquito population (*E*_*v*_), and (c) asymptomatic infectious malaria cases resulting from sub-optimum treatment (*I*_*sa*_), with respect to the parameters of the model (2.1). The indices displayed are calculated at time, *t* = 40. Only the sensitivities of parameters that are significantly different from the dummy variable (*d*) with a p-value less than 0.05 are displayed.

The exposed mosquito population (*E*_*v*_) exhibits similar first order sensitivities, with three additional parameters: the mosquito recruitement rate (Λ_*v*_), the asymptomatic human infectivity modification factor (*θ*), and the fraction of infectious humans who complete malaria treatment (*ε*). The parameters *p*_*vh*_, *µ*_*v*_, and *p*_*hv*_ are the most significant in both first and total order sensitivities. These are followed by *p*_*hv*_, Λ_*v*_, the sub-optimum treatment rate (*ϕ*_*wc*_), the immunity waning rates (*ψ*_*sa*_ and *ψ*_*wa*_), and the treatment rate (*γ*_*sc*_) (Fig. 3 (b) & (e)). The high significance of Λ_*v*_ highlights the crucial role of controlling mosquito populations in malaria management. Effective control measures include targeting mosquito larvae in breeding sites (larviciding), eliminating standing water where mosquitoes (source reduction), and introducing natural predators of mosquito larvae (biological control).

The most influential parameters for asymptomatic infections from sub-optimum treatment, based on both firstorder and total sensitivity indices, are the *p*_*vh*_, *µ*_*v*_, *γ*_*sc*_, *ϕ*_*wc*_, and *ϕ*_*h*1_), followed by *σ*_*v*_, *σ*_*sc*_, *β*_*vh*_, *p*_*hv*_ (Fig. 3(c) and (f)). Although treatment-seeking rates and drug efficacy exhibit no direct significant impact on the outputs, they influence *γ*_*sc*_, *ϕ*_*sc*_, and *ϕ*_*wc*_, which have high sensitivity indices.

In summary, eFAST is used to quantify the impact of model parameters on the exposed human and mosquito populations, and asymptomatic cases. The results show that *p*_*vh*_ has the highest sensitivity index for the three response functions (comparing Fig. 3 (a)-(f)). Also, a small subset of biological and intervention-related parameters drive variability in all three outcomes consistently, underscoring their importance for targeted control. Specifically, public health efforts that prioritize these parameters, particularly those governing transmission and treatment quality are likely to yield the greatest impact. Similar results are obtained for Nigeria (Section 5 of the SI).

#### 3.3.2. Assessing the combined impact of parameters on the reproduction number

Using the parameter values for Kenya in Tables 3-4, contour plots (depicted in Fig. 4) are generated to assess the combined effects of parameter pairs on the reproduction number (ℛ_0_). These include: (a) the rate at which mosquitoes bite infectious humans (*β*_*hv*_) and the treatment rate (*ϕ*_*h*1_); (b) *β*_*hv*_ and the reciprocal of the average time to asymptomatic infection among untreated individuals (*ϕ*_*h*2_); (c) *β*_*hv*_ and the proportion of infectious humans receiving optimum treatment (*ε*); (d) the transition rates *ψ*_*sa*_ (from *I*_*sa*_ → *I*_*sc*_) and *ψ*_*wa*_ (from *I*_*wa*_ → *I*_*wc*_); (e) *β*_*hv*_ and drug resistance, where drug resistance is incorporated by scaling the baseline treatment rate *τ* by the factor (1 − *d*_*r*_), with 0 ≤ *d*_*r*_ ≤ 1; (f) the rate at which mosquitoes bite susceptible humans (*β*_*vh*_) and the mortality rate of mosquitoes (*µ*_*v*_); (g) *β*_*hv*_ and *µ*_*v*_; (h) the mosquito recruitment and mortality rates (Λ_*v*_, *µ*_*v*_); and (i) the asymptomatic human infectivity scaling factor (*θ*) and *µ*_*v*_.

**Figure 4.**
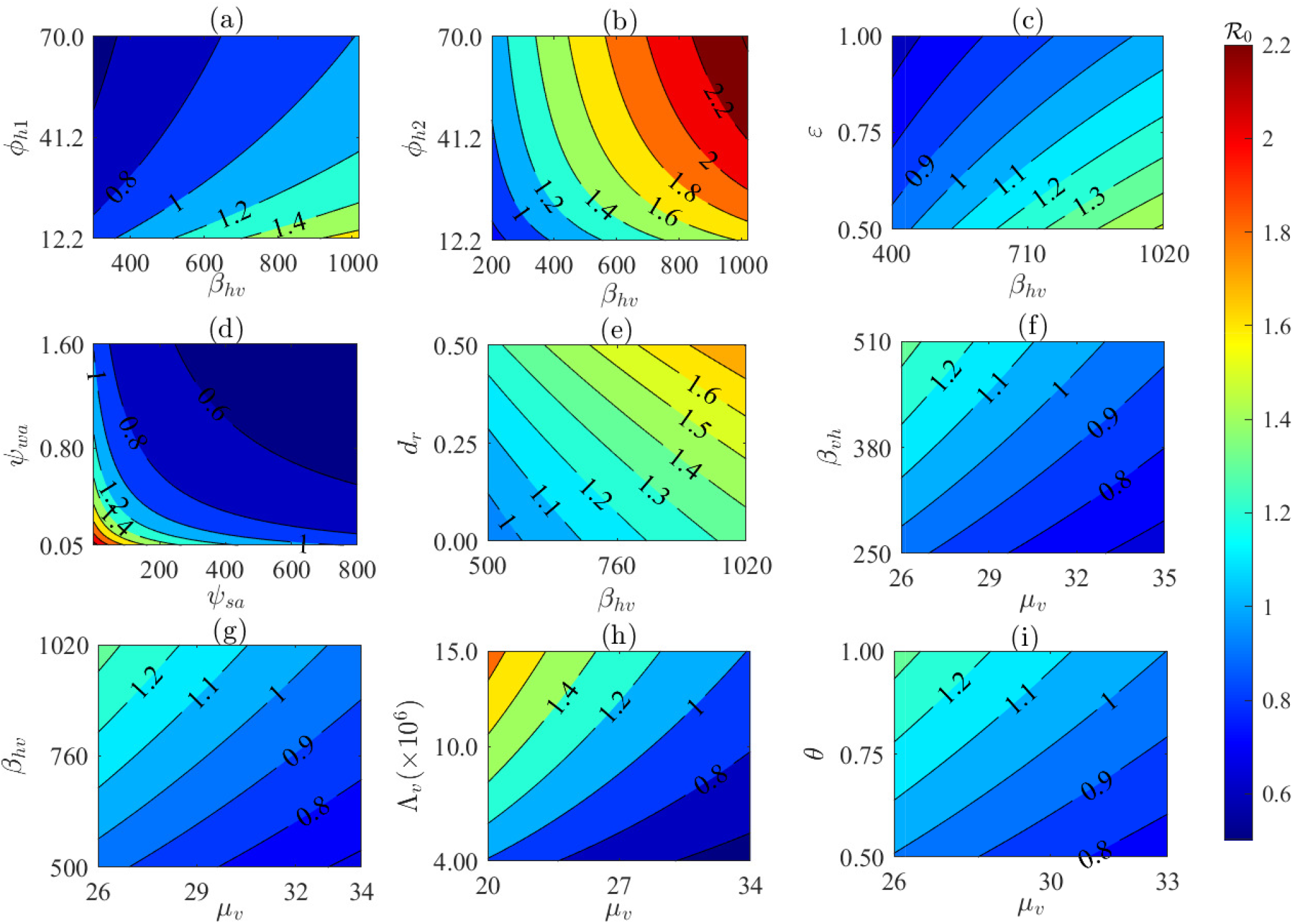
Contour plots showing the impact of pairs of parameters on the reproduction number (ℛ_0_) using the Kenya parameter regime in Tables 3–4. The panels illustrate the joint effects of: (a) *β*_*hv*_ (rate at which mosquitoes bite infectious humans) and *ϕ*_*h*1_ (the treatment rate); (b) *β*_*hv*_ and the reciprocal of the average time to asymptomatic infection among untreated individuals (*ϕ*_*h*2_); (c) *β*_*hv*_ and the proportion of infectious humans receiving optimum treatment (*ε*); (d) the transition rates *ψ*_*sa*_ (from *I*_*sa*_ → *I*_*sc*_) and *ψ*_*wa*_ (from *I*_*wa*_ → *I*_*wc*_); (e) *β*_*hv*_ and drug resistance, modeled by scaling the treatment rate (*τ* ) by (1 − *d*_*r*_), where 0 ≤ *d*_*r*_ ≤ 1; (f) *β*_*vh*_ (the mosquito biting rate on susceptible humans) and the mosquito death rate *µ*_*v*_; (g) *β*_*hv*_ and *µ*_*v*_; (h) the mosquito recruitment and mortality rates (Λ_*v*_, *µ*_*v*_); and (i) the asymptomatic human infectivity factor (*θ*) and *µ*_*v*_.

The results show that malaria control remains achievable in settings with relatively low mosquito biting rates, especially when more individuals seek timely and effective treatment. For example, when the biting rate (*β*_*hv*_) is ≈ 300, halving the treatment duration for first time infections (1/*ϕ*_*h*1_), i.e., setting *ϕ*_*h*1_ ≈ 12.16 is sufficient achieve ℛ_0_ < 1 (Fig. 4 (a)). Even at higher biting rates, control remains possible if *ϕ*_*h*1_ increases sufficiently. However, once *β*_*hv*_ exceeds a critical threshold, reducing ℛ_0_ below unity becomes impossible, making control unattainable regardless of *ϕ*_*h*1_. A similar trend is observed for the treatment rate (*ϕ*_*h*2_), where combinations of *β*_*hv*_ and *ϕ*_*h*2_ can bring ℛ_0_ below one, but control becomes unlikely beyond a certain biting rate (Fig. 4 (b)). Thus, in settings like Kenya, where some individuals do not seek treatment and become asymptomatic carriers, malaria can only be contained when mosquito biting rates remain below some critical threshold. The results for the case of *β*_*hv*_ and *ε* emphasize the role of treatment completion in curtailing malaria. Specifically, higher rates of incomplete treatment require substantially lower biting rates for control and vice versa (Fig. 4 (c)). In particular, if 67% of infectious humans fail to complete treatment (i.e., *ε* = 0.67), control is possible if *β*_*hv*_ ≈ 581, whereas if 88% do not complete treatment, reducing ℛ_0_ below one requires a higher biting rate of about 804.6. Once *β*_*hv*_ ≥ 954, reducing ℛ_0_ below one is impossible and hence, malaria persists even with perfect treatment completion.

The waiting times (1/*ψ*_*sa*_ and 1/*ψ*_*wa*_) in the asymptomatic compartments (*I*_*sa*_ and *I*_*wa*_) before progression to the symptomatic infection compartments (*I*_*sc*_ and *I*_*wc*_) are key determinants of transmission. In particular, shorter waiting periods result in higher transition rates (*ψ*_*sa*_ and *ψ*_*wa*_) and a reduction in ℛ_0_, whereas longer waiting periods result in lower transition rates and hence an increase in ℛ_0_ (Fig. 4(d)). For a short waiting time in *I*_*sa*_ (e.g., 1/*ψ*_*sa*_ = 0.0059, i.e., *ψ*_*sa*_ = 169.73), a waiting time of 1/*ψ*_*wa*_ = 3.0139 (i.e., *ψ*_*wa*_ = 0.3318) in *I*_*wa*_ is sufficient to bring ℛ_0_ below one. For a longer waiting time (e.g., 1/*ψ*_*sa*_ = 0.0112, i.e. *ψ*_*sa*_ = 88.9333), a waiting time of 1/*ψ*_*wa*_ = 1.7649 (i.e., *ψ*_*wa*_ = 0.5666) in *I*_*wa*_ is enough to ensure control of transmission. A similar pattern is observed when the roles of the two asymptomatic compartments are reversed, illustrating the trade-off between the stages in determining overall transmissibility. Results assessing the impact of mosquito biting and drug resistance on ℛ_0_ (Fig. 4(e)) show that high drug resistance combined with high mosquito biting rates renders ℛ_0_ ≤ 1 unattainable, while reducing ℛ_0_ below one is possible with low resistance and low mosquito biting. In particular, without resistance (i.e., *d*_*r*_ = 0), ℛ_0_ < 1 is achievable when *β*_*hv*_ ≤ 563 *per year*, but reducing ℛ_0_ below one is impossible even with the same biting rate once resistance emerges (i.e., *d*_*r*_ = 0), except both the level of resistance and the biting rate are very low. Reductions in mosquito biting rates and increases in mosquito mortality play a key role in lowering ℛ_0_. For example, if the baseline biting rates (*β*_*vh*_ and *β*_*hv*_) are reduced by 15% while the baseline mosquito mortality rate (*µ*_*v*_) is increased by 15%, achieving ℛ_0_ < 1 remains impossible (Fig. 4(f) and (g)). However, if the baseline biting rate is reduced by 20% and the baseline mosquito mortality rate is increased by 20%, then ℛ_0_ < 1 becomes achievable (Fig. 4(f) and (g)). This illustrates the complementary roles of lowering mosquito–human contact and shortening mosquito lifespan in reducing transmission potential. In practice, reductions in biting can be achieved through interventions such as ITNs, repellents, and protective clothing, while increases in adult mosquito mortality can be promoted through ITNs, and IRS. The contour plots also show that reducing mosquito population growth and survival (i.e., reducing mosquito recruitment) and increasing mosquito mortality simultaneously are important measures for attaining ℛ_0_ < 1. Such interventions include larviciding, removal of breeding sites, IRS, and ITN-use (Fig. 4 (h)). Specifically, if Λ_*v*_ ≤ 4 million *mosquitoes per year* and the average lifespan of mosquitoes is 19.7 *days*, (i.e., *µ*_*v*_ ≤ 20 *per year*), ℛ_0_ cannot exceed unity, but if Λ_*v*_ = 10^7^ *mosquitoes per year* and the average lifespan of mosquitoes is 13 *days*, (i.e., *µ*_*v*_ ≤ 28 *per year*), ℛ_0_ > 1. Figure 4 indicates that when the asymptomatic transmission parameter *θ* ≤ 75%, increasing *µ*_*v*_ up to 30 *per day* is sufficient to achieve ℛ_0_ < 1. Higher values of *θ* require proportionally larger *µ*_*v*_; for example, *θ* = 96% necessitates *µ*_*v*_ ≥ 32.6 *per day* to ensure ℛ_0_ < 1.

The results highlight key public health strategies for malaria control. In particular, effective malaria control requires integrated strategies that simultaneously reduce human–mosquito contact and strengthen case management. While timely treatment, high treatment completion, and faster progression from asymptomatic to symptomatic infection reduce transmission, these measures alone are insufficient when mosquito biting rates exceed critical thresholds. Complementary vector control measures such as ITNs, IRS, larval source management, and environmental control are therefore essential to lower biting rates, increase mosquito mortality, limit vector recruitment, and ultimately drive ℛ_0_ below unity. Comparable simulations using the Nigerian parameter regime (Tables 3–4, Fig. 2 in the SI) indicate that malaria is more likely to be contained in Kenya than in Nigeria, consistent with higher estimated reproduction numbers in the latter (Table 4).

#### 3.3.3. Assessing the impact of mosquito biting rates and infection probabilities

We assessed how changes in the transmission probabilities from mosquitoes to humans (*p*_*vh*_) and from infectious humans to mosquitoes (*p*_*hv*_) shape malaria dynamics by fixing one probability at its fitted value and increasing or decreasing the other, or both, by 25% (Fig. 5(a)-(b), (e)-(f)). Specifically, holding *p*_*vh*_ at its baseline value, a 25% increase in *p*_*hv*_ raises the peak number of symptomatic infectious humans by approximately 36% (blue vs. red curves in Fig. 5(a)). Conversely, increasing *p*_*vh*_ by 25% while keeping *p*_*hv*_ at baseline produces a 30% rise (blue vs. green curves). When both transmission probabilities increase by 25%, the peak increases most substantially, by about 68% (blue vs. yellow curves). On the other hand, reducing *p*_*hv*_ by 25% while maintaining *p*_*vh*_ at its baseline value lowers the the peak number of symptomatic infectious humans by about 42% (blue vs. red curves in Fig. 5(b)). Reducing *p*_*vh*_ by the same proportion while keeping *p*_*hv*_ at baseline leads to a 39% decrease (blue vs. green curves in Fig. 5(b)), and a simultaneous 25% reduction in both probabilities produces the largest reduction of 78% (blue vs. yellow curves in Fig. 5(b)). Similar qualitative changes are observed with the the peak number of symptomatic infectious humans cumulative number of confirmed cases (Fig. 5(e)-(f)). A consistent pattern emerges when comparing changes in *p*_*vh*_ and *p*_*hv*_. Specifically, increasing the human-to-mosquito transmission probability (*p*_*hv*_) has a greater impact on epidemic peaks and cumulative cases than an equivalent increase in the mosquito-to-human infection probability (*p*_*vh*_), and the same asymmetry appears for decreases.

**Figure 5.**
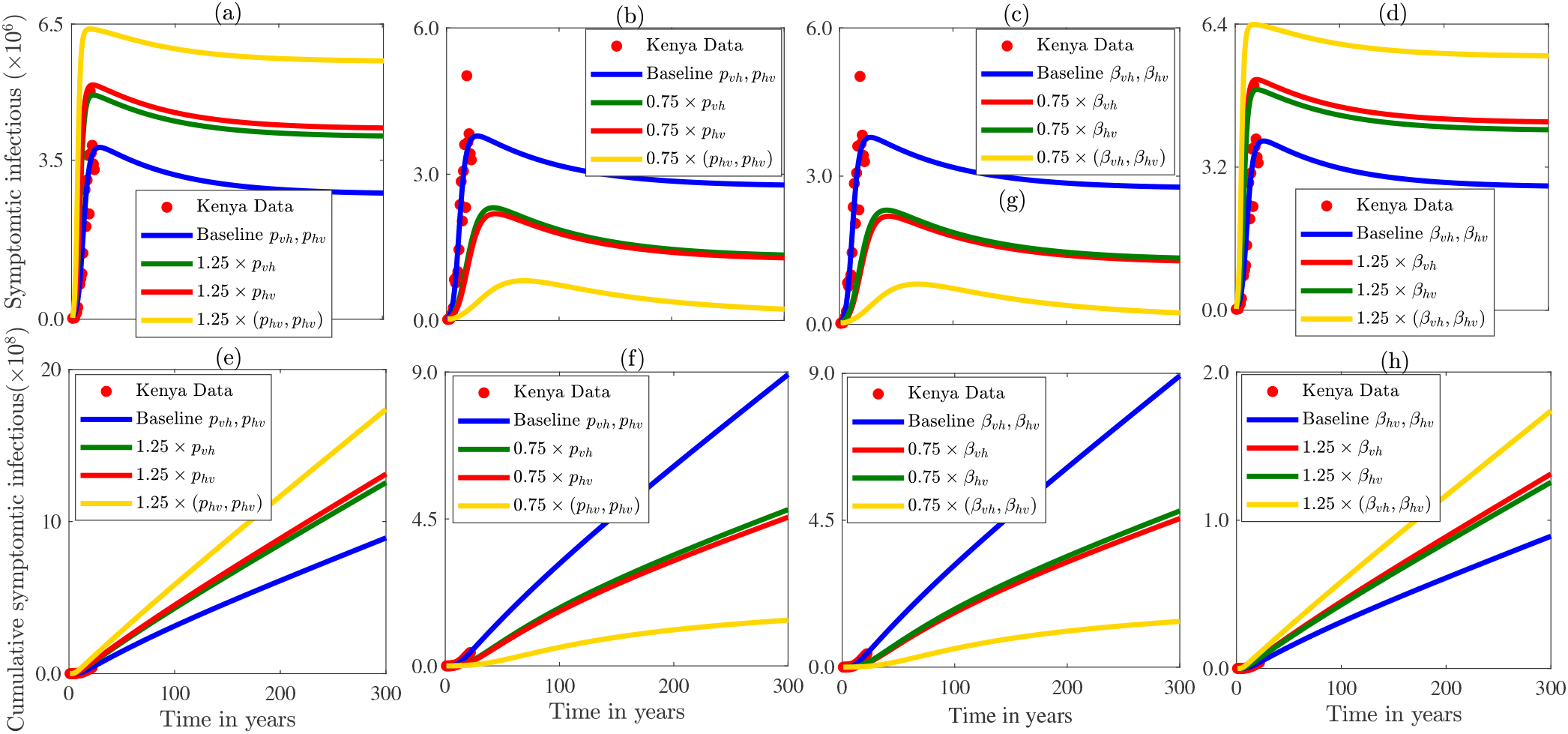
Illustration of the effects of varying the human-to-mosquito transmission probability (*p*_*hv*_), the mosquito-to-human transmission probability (*p*_*vh*_) (panels (a)–(b)), and the mosquito biting rates *β*_*vh*_ and *β*_*hv*_ (panel (c)-(d)) on the annual number of exposed humans. Panels (e)–(h) show the corresponding cumulative exposed populations. All other parameter values used in the simulations are for Kenya and presented in Tables 3 and 4.

Also, we examined how heterogeneity in the mosquito biting rates on susceptible humans (*β*_*vh*_) and on infectious humans (*β*_*hv*_) influences malaria dynamics by considering scenarios in which one biting rate is kept at its baseline value while the other is increased or reduced by 25%, as well as the case in which the two rates are reduced or increased by 25% (Fig. 5(c)-(d) & (g)-(h) ). When *β*_*hv*_ is fixed at its baseline value, reducing *β*_*vh*_ by 25% lowers the peak number of symptomatic infectious humans by approximately 42% (blue vs. red curves in Fig. 5(c)), whereas if *β*_*vh*_ is fixed at its baseline value, reducing *β*_*hv*_ by 25% lowers the peak number of symptomatic infectious humans by approximately 39% (blue vs. green curves in Fig. 5(c)). Reducing both *β*_*hv*_ and *β*_*vh*_ by 25% lowers the peak size by about 78% relative to the baseline (yellow vs. blue curves in Fig. 5(c)). In contrast, if both *β*_*vh*_ and *β*_*hv*_ increase by 25%, the resulting peak is roughly twice that observed when only *β*_*vh*_ increases (Fig. 5(d)). When *β*_*hv*_ is fixed at its baseline value, increasing *β*_*vh*_ by 25% raises the peak number of symptomatic infectious humans by approximately 36% (blue vs. red curves in Fig. 5(d)). Similar reductions and increases are reflected in the cumulative number of symptomatic infectious humans (Fig. 5(g)–(h)). These results show that the peak size is more sensitive *β*_*hv*_ than *β*_*vh*_, mirroring the asymmetry observed for *p*_*vh*_ and *p*_*hv*_. Thus, Increased biting activity on infectious humans enhances transmission more than increased in biting on susceptible individuals, while reductions in biting activities have a larger mitigating effect. Strategies that reduce both *β*_*vh*_ and *β*_*hv*_ remain the most effective. Similar results using the parameters for Nigeria are presented in Fig. 4 of the SI.

#### 3.3.4. Assessing the impact of optimum and sub-optimum treatment on malaria dynamics

The model (2.1) is simulated using the parameter values for Kenya in Tables 3 and 4 to investigate how optimum and sub-optimum treatment shape malaria transmission dynamics. Three parameters were used to investigate this phenomenon–the proportion of symptomatic infectious humans who seek treatment, *p*, the proportion (*ε*) of infectious humans, who complete treatment (optimum treatment), and the sub-optimum treatment rate *ϕ*_*wc*_. The results show that incomplete treatment, especially infectious individuals who stop treatment increases malaria transmission significantly. Halving the baseline value of *p* leads to a 111% increase in the baseline peak number of symptomatic cases (blue versus red curves in Fig. 6(a)). Increasing *p* by 50% or 200% reduces the peak by 51% and 97%, respectively (blue versus magenta and green curves in Fig. 6(a). These results indicate that improving treatment-seeking has substantial public-health benefits: increases in *p* reduce both peak and cumulative symptomatic cases (Fig. 6(a) & (d)), and analogous outcomes using Nigeria-specific parameter values are presented in Fig. 3 of the SI.

**Figure 6.**
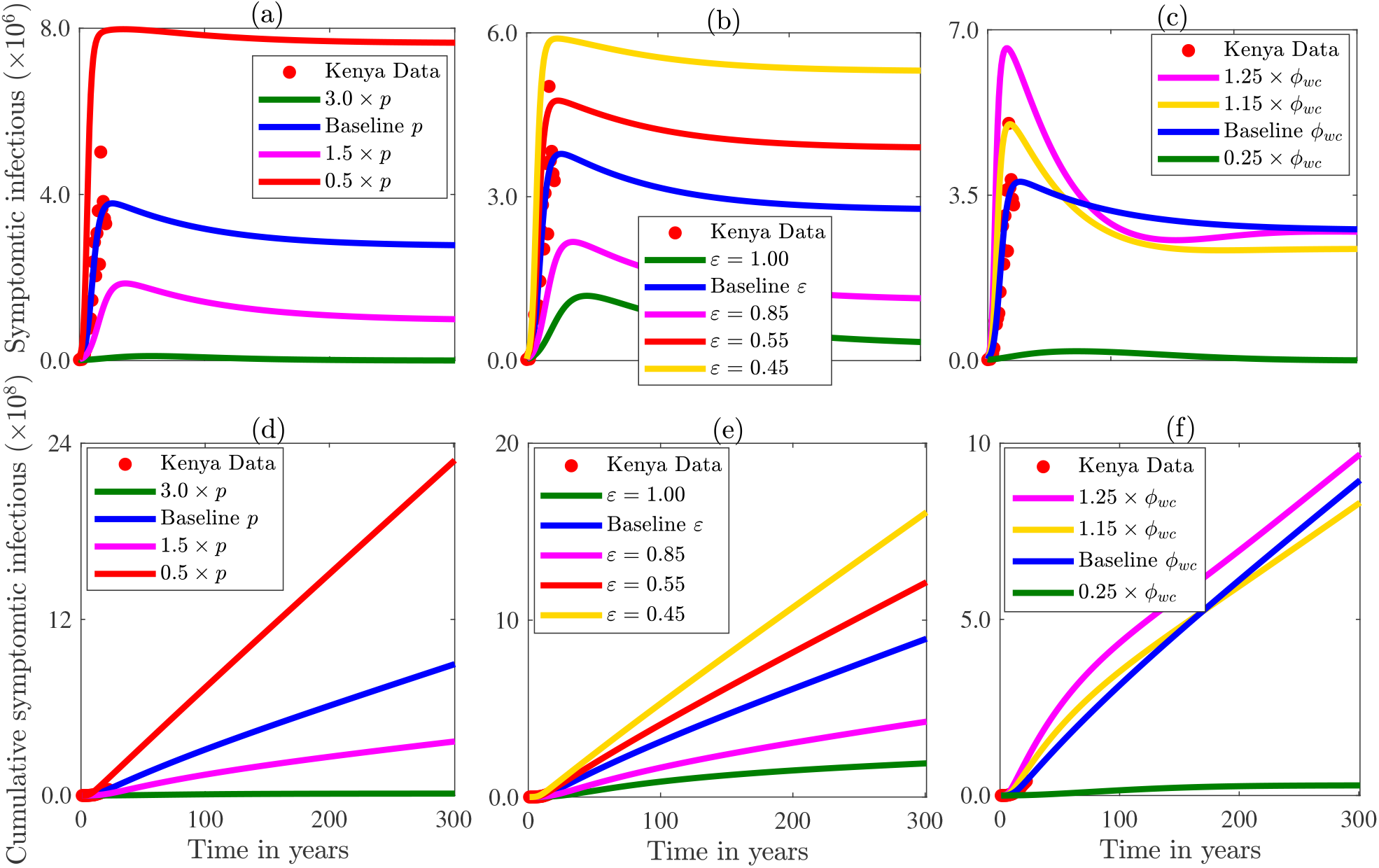
Simulations of Model (2.1) showing the impact on the yearly number of symptomatic infectious cases of the (a) fraction of humans who seek treatment (*p*), (b) fraction of humans who complete treatment (*ε*), and (c) the sub-optimum treatment rate (*ϕ*_*wc*_). Corresponding cumulative results are shown in (d)-(f). The other parameter values used for the simulations are given in Tables 3-4.

Improper or incomplete treatment remains a major obstacle to malaria control, largely because it accelerates the transition from symptomatic to asymptomatic infection. This effect is captured by the sub-optimum treatment rate *ϕ*_*sc*_ (*ϕ*_*wc*_) for the *I*_*sc*_ (*I*_*wc*_) class, while optimum treatment is accounted for by the proportion of individuals who receive optimum treatment (*ε*). To assess the impact of treatment completion, we consider four values of *ε*: 1, 0.85, 0.55, and 0.45. Raising *ε* to 1 (i.e., every infectious individual receives complete treatment) or 0.85 (i.e., 85% of infectious individuals receive optimum treatment) reduces the baseline peak number of symptomatic infectious by ≈ 69% and ≈ 42% (green and magenta versus blue curves in Fig. 6(b)). Conversely, lowering treatment completion worsens transmission. In particular, decreasing *ε* by 10% (to 0.55) or by 20% (to 0.45) increases the baseline peak number of symptomatic infectious by over 26% or 56%, respectively (red and yellow versus blue curves in Fig. 6(b)). Figure 6(c) shows that increases in the sub-optimum treatment rate *ϕ*_*wc*_ result in more transmission and hence an increase in the baseline number of confirmed cases. In particular, a 25% rise in this rate results in a 75% increase in the baseline peak number of yearly symptomatic infectious (magenta versus blue curve in Fig. 6(c)). By contrast, reducing *ϕ*_*wc*_ by 75% markedly improves control (green versus blue curve in Fig. 6(c)). These results underscore the substantial public health gains achievable through improved treatment adherence: More optimum and less sub-optimum treatment consistently lower both the peak and cumulative number of confirmed cases. Analogous results generated using parameter values for Nigeria are presented in Fig. 3 of the SI.

#### 3.3.5. Assessing the impact of asymptomatic transmission and sub-optimum treatment on malaria burden

To quantify the contribution of asymptomatic infections to the burden of malaria in high-endemic settings with variable treatment efficacy, we use Disability-Adjusted Life Years (DALYs). DALYs combine years of life lost due to premature mortality (YLL) with years lived with disability (YLD), providing an integrated measure of both fatal and non-fatal disease outcomes. Following world health organization methodology,

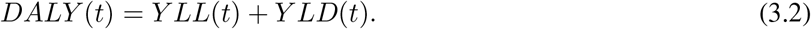

Here, *Y LL*(*t*) = *η* × *L*, where *η* is the number of malaria deaths and *L* is the remaining life expectancy at age of death. The disability component is given by *Y LD* = *DW* × *γ* × *I*, where *DW* is the disability weight, *I* is confirmed cases, and *γ* is the average duration of infection. Treated individuals experience a mean disease duration of ≈ 14 days, whereas untreated cases progress to an asymptomatic state after ≈ 60 days. Disability weights vary by severity: 0.006 (mild), 0.051 (moderate), 0.133 (severe), and 0.219 (post-acute) [55], necessitating separate YLD estimates for symptomatic and asymptomatic humans. Because disease deaths are in the symptomatic class, *Y LL* is computed for this class, while quantify the asymptomatic contribution to *Y LD* (full national DALY values are found in [56]. The disability weights for symptomatic and asymptomatic humans denoted by *DW*_*s*_ and *DW*_*a*_, respectively, with *DW*_*a*_ ∈ {0.006, 0.012, 0.018} and *DW*_*s*_ ∈ {0.219, 0.133, 0.051}, are used to compute *Y LD*.

The DALY-derived *Y LD* estimates for Kenya (top row) and Nigeria (bottom row) are shown in Fig. 7(a)–(f). Panels (a) and (d) show total *Y LD* under three disability-weight combinations; panels (b) and (e) display the symptomatic contribution; and panels (c) and (f) show the asymptomatic contribution for (*DW*_*a*_, *DW*_*s*_) = (0.006, 0.219), with green and blue curves illustrating two-and three-fold increases in *DW*_*a*_. Across all scenarios, *Y LD* increases with the disability weights as expected. For Kenya, the highest burden arises when (*DW*_*a*_, *DW*_*s*_) = (0.006, 0.219) (green curves), followed by (0.006, 0.133) and (0.006, 0.051) (blue and magenta curves). Specifically, in 2021 the largest estimated *Y LD* is roughly 1.12 times the smallest, and the intermediate weight yields values ≈ 6% higher than the lowest. Although symptomatic infections dominate *Y LD* under baseline weights, this reflects their larger *DW*_*s*_ rather than their numerical prevalence: increasing *DW*_*a*_ to 0.018 raises asymptomatic *Y LD* to nearly 80 times the symptomatic contribution computed using *DW*_*s*_ = 0.051 (comparing magenta curves in Fig. 7(b)-(c)). The burden for Nigeria shows the same qualitative pattern but with a higher overall burden (more than twice that for Kenya). Moreover, asymptomatic infections contribute a larger fraction of *Y LD* in Kenya. In particular, in 2022, asymptomatic infections account for ≈ 96% of Kenya’s total *Y LD* but only ≈ 75% of Nigeria’s. This aligns with documented disparities in treatment access and policies. In Kenya, successive changes in drug guidelines, declining ACT subsidies, and resistance reduced treatment effectiveness (resulting in increased asymptomatic carriers), whereas Nigeria maintained consistent treatment practices over the same period [49, 57–59].

**Figure 7.**
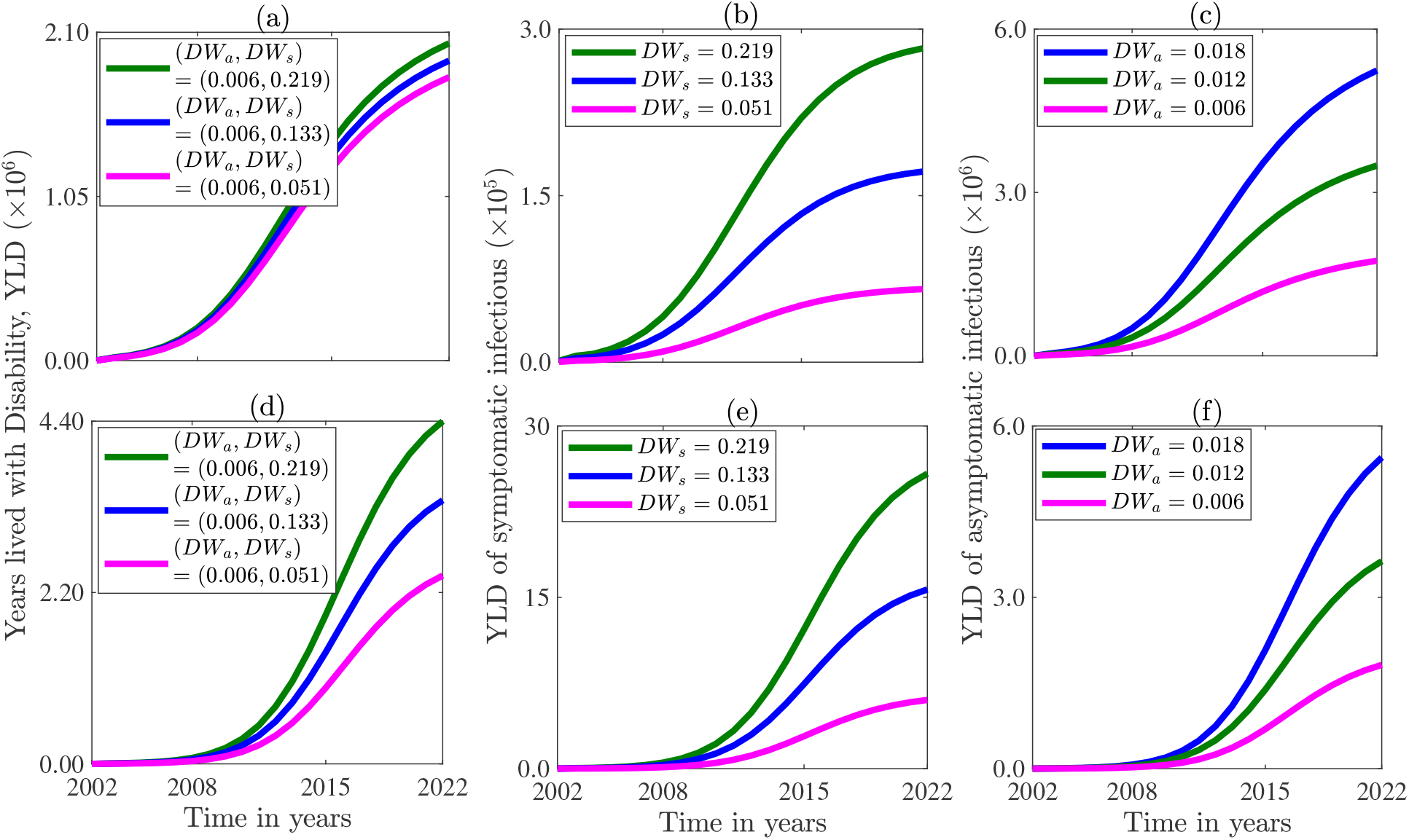
Simulations of the model (2.1) depicting Years Lived with Disability (YLD) ((a) and (d)) and the proportional contributions of symptomatic infections ((b) and (e)) and asymptomatic infections ((c) and (f)) for Kenya ((a)-(c)) and Nigeria ((d)-(f)) from 2002–2022 under three disability-weight combinations. The parameter values used are presented in Tables 3–4.

To further contextualize these patterns, we compare the proportional contributions of *Y LD* and total DALYs under alternative disability-weights relative (*DW*_*a*_, *DW*_*s*_) = (0.006, 0.219) early, mid-, and late stages in our model fitting window. For each country, increases in *DW*_*a*_ or decreases in *DW*_*s*_ lead to increases in the share of *Y LD* attributable to asymptomatic infections. For (*DW*_*a*_, *DW*_*s*_) = (0.012, 0.133), asymptomatic *Y LD* rises to approximately ≈ 100% (early, mid-, and late) of the baseline values for Kenya, and Nigeria. The increases are more pronounced, reaching ≈ 200%, of the baseline for Kenya and Nigeria when (*DW*_*a*_, *DW*_*s*_) = (0.018, 0.051) (comparing yellow and magenta curves in Fig. 7(b)–(c)). When translated into DALYs, the same trend is observed; scenarios with high *DW*_*a*_ shift a larger fraction of the total disease burden toward asymptomatic infections, with asymptomatic DALYs exceeding the baseline proportions by ≈ 100%–200% for Kenya and Nigeria across the three time points. These comparisons underscore the importance of characterizing asymptomatic disability weights, as modest changes in *DW*_*a*_ alter the burden and resource-allocation priorities within both countries significantly.

#### 3.3.6. Optimum versus sub-optimum treatment costs

White *et al*. [19] carried a broad cost review of malaria interventions that covered vector control, preventive measures, diagnostics, and treatment. This review that focused primarily on sub-Saharan African studies (78%), reported median financial costs ranging from $5.84 to $30.26 for treating uncomplicated to severe malaria, with a median diagnostic cost of $4.32 per case. Incorporating diagnostic costs yields per-person treatment costs of $10.16 for uncomplicated and $34.58 for severe malaria. To quantify the financial burden of malaria treatment, we estimate costs by multiplying the unit diagnostic and case-management costs from White et al. [19] with the numbers of symptomatic and asymptomatic cases from our model. Figure 8 shows the total cost of treatment for both asymptomatic and symptomatic infectious humans (red curve in Fig. 8), treating asymptomatic infections only (magenta curve in Fig. 8), and treating symptomatic infections only (blue curve in Fig. 8). A comparison of optimum and sub-optimum treatment costs are depicted in Fig. 8(c)-(d). Additional expenditures due to sub-optimum treatment estimated at the peak time for Kenya (Nigeria) is ≈ 1.69, (5) times higher than the optimum treatment cost. These additional costs are even higher near the equilibrium indicating that sub-optimum treatment increases short-and long-term costs. Lower costs of more than 60% are achievable with optimum treatment.

**Figure 8.**
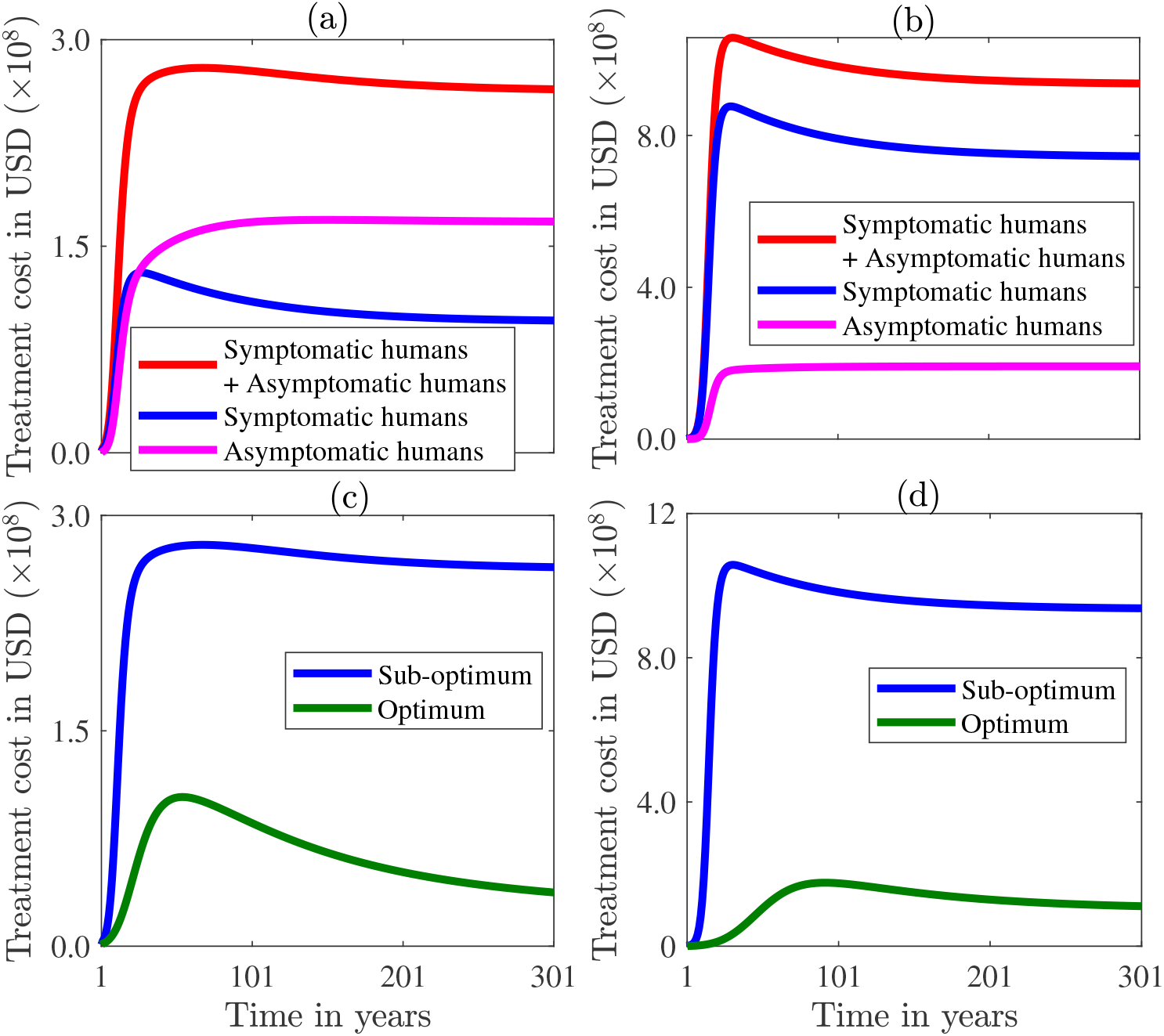
Estimated cost of malaria treatment. (a)–(b) Sub-optimum treatment costs for symptomatic, asymptomatic, and combined symptomatic and asymptomatic infectious humans for (a) Kenya and (b) Nigeria. (c)–(d) Comparison of optimum and sub-optimum treatment costs for (c) Kenya and (d) Nigeria. The parameter values used are given in Tables 3 and 4.

The treatment cost of asymptomatic (symptomatic) malaria is estimated using the cost of uncomplicated (severe) cases due to the mild (severe) symptoms. Our simulations indicate that, at the endemic equilibrium, the number of asymptomatic (symptomatic) cases are ≈ 1.65 × 10^7^ (2.75 × 10^6^), yielding a ratio of ≈ 6 asymptomatic cases per symptomatic case. In a hypothetical scenario with full optimum treatment (i.e., *ε* = 1), the number of asymptomatic cases reduces by ≈ 33% (50%) for Kenya (Nigeria). Since *I*_*sa*_ represents individuals who become asymptomatic following sub-optimum treatment and subsequently progress to symptomatic infection at rate *ψ*_*sa*_, the additional symptomatic cases requiring treatment at equilibrium are estimated as *ψ*_*sa*_ times the difference between the baseline level of *I*_*sa*_ and its level under optimum treatment. This corresponds to 347,220 additional symptomatic cases and an estimated annual treatment cost of $12,007,000 for Kenya attributable to prior sub-optimum treatment.

## 4. Discussion, limitations, and concluding remarks

### 4.1. Discussion

Malaria, a mosquito-borne communicable disease transmitted by female *Anopheles* mosquitoes [60, 61], remains endemic especially in sub-Saharan Africa despite extensive and long-standing pharmaceutical and vector-control measures. Core intervention strategies include the use of ITNs, larviciding, and IRS. Although two vaccines, RTS,S/AS01 (2021) and R21/Matrix-M (2023) [58], have been approved, their moderate efficacies in preventing symptomatic infection (≈ 75% for RTS,S/AS01 and 55% for R21/Matrix-M) and their current restriction to children mean that antimalarial drugs and vector control remain core control measures. When administered properly, drugs can clear infections with up to 95% efficacy [62]. In malaria-endemic regions, however, treatment is frequently sub-optimum due financial barriers, drug and insecticide resistance, negligence, and sociocultural factors [63]. These challenges contribute to a substantial, often undetected reservoir of asymptomatic infections that complicate surveillance, sustains transmission, and undermines the effectiveness of control measures. Understanding the epidemiological and economic consequences of sub-optimum treatment is therefore critical for designing effective intervention strategies. In this study, we develop and use a model to investigate how sub-optimum treatment sustains asymptomatic infections, quantify the resulting economic burden relative to optimum treatment, and evaluate implications for malaria control. The model captures treatment-induced progression pathways and preferential mosquito feeding on infectious hosts. It is parameterized using yearly case data from Kenya and Nigeria.

Fitting the model (2.1) to data, shows that sub-optimum treatment sustains asymptomatic infections and increases long-term transmission. The estimated parameters reveal epidemiological contrasts between Kenya and Nigeria. The reproduction number for each setting confirm endemic transmission and show more intense spread in Nigeria, aligning with WHO assessments [64]. Furthermore, the mosquito to human transmission probability (*p*_*vh*_) exceeds that from humans to mosquitoes (*p*_*hv*_), which is consistent with earlier work [51]. These findings emphasize the importance of treatment completion and strengthening context-specific control strategies in endemic regions [65].

A global uncertainty and sensitivity analysis using eFAST [26, 52, 54], which is well-suited for nonlinear systems such as the model (2.1) shows that the pathogen transmission from mosquitoes to humans, mosquito demographic and biting parameters consistently have the greatest influence on model outputs, aligning with evidence that vector survival and feeding behavior dominate malaria transmission dynamics [26]. The high impact of mosquito mortality and biting rates reinforces the importance of interventions that reduce vector populations and human–mosquito contact, e.g., ITNs, IRS, larval source management, and complementary community based measures [26]. These results situate our findings within the broader malaria control literature by confirming that vector targeted strategies remain the most powerful measures for reducing transmission in endemic settings such as Kenya and Nigeria. Also, they emphasize the need for sustained investment in interventions that curtail mosquito survival and biting to achieve meaningful and lasting public health gains. Sensitivity of some of the response functions to the sub-optimum treatment rate and mosquito birth rate underscores the dual importance of strengthening treatment adherence and reducing vector recruitment. These findings align with studies showing that environmental management, larval source reduction, larviciding, biological control, and emerging genetic mosquito-suppression tools can reduce vector populations substantially and disrupt transmission [26]. Together with our model-based evidence from Kenya and Nigeria, these results reinforce the fact that sustained investment in interventions that improve optimum treatment and reduce mosquito survival are central to reducing malaria and advancing long-term control.

Local sensitivity analysis of the model (2.1) shows that malaria dynamics are most sensitive to parameters that are related to mosquito-to-human transmission, with increases in mosquito infectivity (*p*_*vh*_) and biting rate on susceptible hosts (*β*_*vh*_) triggering greater increases in malaria case counts than equivalent changes in human-to-mosquito infectivity or the rate at which mosquitoes bite infectious individuals. This is consistent with longstanding evidence that human risk is driven primarily by vector infectivity and biting pressure [66]. These results reinforce the dominant role of interventions that reduce mosquito-to-human transmission, such as ITNs, IRS, larval source management, repellents, and environmental strategies that limit exposure [26, 67]. Treatment-related parameters are also highly sensitive to the number of malaria cases. In particular, decreases in treatment seeking (*p*), lower treatment completion (*ε*), and increases in sub-optimum treatment rates (*ϕ*_*sc*_, *ϕ*_*wc*_) lead to higher peak case sizes and cumulative infections in both Kenya and Nigeria. The sensitivity patterns show that malaria burden responds strongly to reductions in mosquito infectivity and biting pressure, but that strengthening treatment adherence provides an essential complementary pathway for curbing transmission, supporting integrated vector-control and case-management strategies as the most effective pathways for sustained control.

An important measure of malaria burden due to the asymptomatic cases is Years lived with Disability, *Y LD* (a critical component of DALYs). Our study shows that asymptomatic carriers contribute up to 96% (75%) of the *Y LD* in Kenya (Nigeria) even with the smallest Disability weight of 0.6%. Thus, failure to account for asymptomatic cases in DALY calculations can underestimate the burden of malaria significantly. The high contribution of asymptomatic cases to *Y LD* and hence to the DALYs in Kenya compared to Nigeria may be explained by the difference in control measures, e.g., anti-malarial access, health infrastructure, and treatment adherence are generally stronger in Kenya than in Nigeria, as reflected in higher ITN-coverage, broader community health worker programs, and comparatively lower malaria prevalence. Our results indicate that incomplete treatment increases asymptomatic infections and raises malaria treatment costs by over $12 million in low-transmission settings, with higher costs in high-transmission settings such as Nigeria. Additional indirect costs, e.g., productivity losses and complications arising when asymptomatic infections relapse to symptomatic infections [68], are not included. Our recommendation to identify and treat asymptomatic individuals to reduce malaria burden and improve control aligns with [69].

Our cost analysis shows that sub-optimum treatment increases the economic burden of malaria substantially. This aligns with prior evidence that inadequate case management drives long-term system-level costs [19]. Simulations using data from Kenya show large symptomatic burdens and substantial financial losses attributable to incomplete treatment, whereas improving treatment completion yields sizeable reductions in both case numbers and overall spending. Notably, eliminating sub-optimum treatment could save more than $12 million annually through fewer secondary symptomatic cases and reduced need for mass treatment, indicating that investments in strengthening diagnostic coverage, adherence support, and high-quality case management are both cost-saving and epidemiologically impactful. These results confirm the fact that effective case management, which can result in immediate health gains and sustained economic benefits in high-burden settings complements vector control in malaria interventions.

### 4.2. Limitations

The model in this study relies on simplifying assumptions. Specifically, the model assumes homogeneous malaria transmission across space because of limited access to county level data. With this assumption the study overlooks spatial heterogeneity in mosquito abundance, environmental conditions, and human exposure that influences malaria risk. Also, the study does not account for the stochastic nature of malaria transmission, despite evidence that random fluctuations in mosquito populations and human behavior can alter transmission dynamics. Likewise, the model omits seasonal variation, even though rainfall and temperature patterns are known to drive the periodic surges in malaria cases observed in endemic regions. Additionally, the model does not incorporate age-dependent transmission, although malaria risk and immune response may vary significantly across age groups. Addressing these limitations in future work would yield a more comprehensive and context-sensitive understanding of malaria dynamics including accounting for heterogeneity across populations through the use of more granular data.

### 4.3. Conclusion

This study uses a mathematical model to show that sub-optimum malaria treatment is a critical yet overlooked driver of persistent transmission and hidden asymptomatic infections. The model that is trained with data from Kenya and Nigeria, is used to show that strengthening adherence to optimum malaria treatment can avert over one-third of infections and deaths, while preventing millions in avoidable economic losses, suggesting that high-quality, complete treatment is substantially beneficial for malaria control. The study identifies demographic, transmission, and treatment related parameters that drive outcome variability consistently, indicating that targeting these key drivers can offer the greatest potential for impactful malaria control. Our findings challenge the conventional focus on vector control, demonstrating that sustainable malaria control requires emphasis on treatment completion, asymptomatic case detection, and integrated vector–treatment strategies. Hence, we conclude that in addition to vector control, public health policies that strengthen treatment adherence and target asymptomatic carriers can reduce transmission, disease burden, and economic cost–shaping a more effective and sustainable control pathway.

## Data Availability

All data produced in the present study are available upon reasonable request to the authors

## Acknowledgement

This work was supported by the National Science Foundation under Grant Numbers DMS-2151870 (CNN, HBT, MS, JER), DMS-2151871 (OP, MP), and DMS-2151872 (RZ). The authors thank Hristina Kostadinova (HK) for her assistance with the initial malaria data compilation and Tuyen Truong (TT) for support related to the model’s endemic equilibrium and reproduction number. Their helpful contributions strengthened the project but do not meet authorship criteria under standard guidelines. All three contributors were supported by DMS-2151870.

