## Supplementary Information for "Unveiling the hidden threat: the impact of sub-optimum treatment on acquired immunity, asymptomatic cases, and malaria dynamics"

### Supplementary Information (SI)

#### 1. Fitting of the net population of Kenya and Nigeria

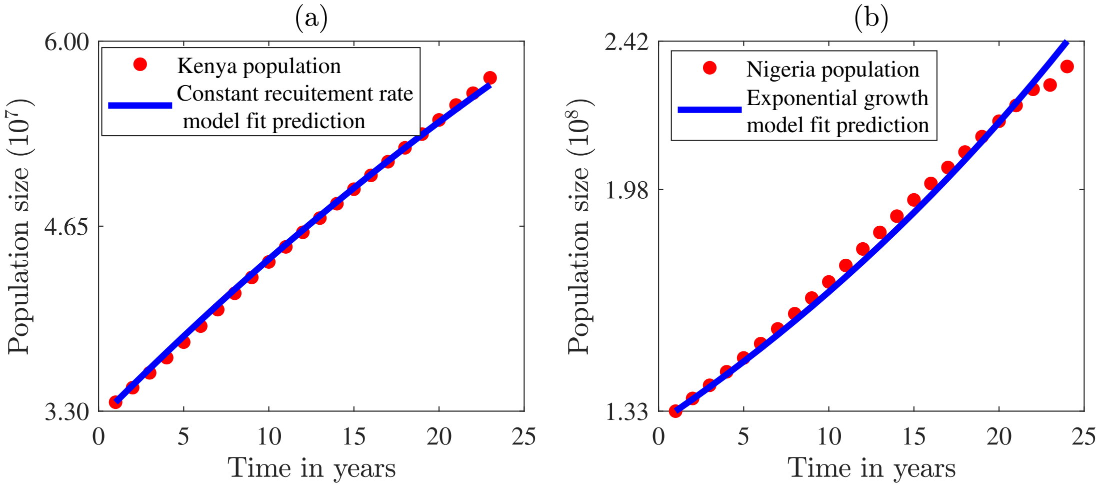


Figure 1: Illustration of the model fits used to estimate the recruitment rates reported in Table 3 of the main paper. (a) A fit of the weak logistic growth model ($\dot{N} = \Lambda - \mu N$), to Kenyan population growth data. (b) Nigeria population An exponential growth model ($\dot{N} = (\eta - \mu)N$) fitted to Nigerian's population growth data. Here, $\Lambda$ is the constant recruitment rate, $\mu$ is the natural death rate of humans and $\eta$ is the birth rate ($\eta - \mu > 0$).

#### 2. Impact of modification of pairs of parameters on dynamics on Malaria in Nigeria

For comparison, and consistent with Figure 4 in the main text, Figure 2 was generated using the Nigeria-specific parameter values reported in Tables 3–4. Figure 2(a) shows that malaria control is achievable under two distinct regimes: either when the mosquito-to-human biting rate on infected individuals ($\beta_{hv}$) is moderately low with a low treatment rate for first-time infections ($\phi_{h1}$), or when both parameters are high. For instance, if $\beta_{hv} \leq 523.232$, then $\phi_{h1} \geq 12.167$ ensures $\mathcal{R}_0 < 1$. Similarly, when $\beta_{hv} \approx 1703.03$, control is feasible provided $\phi_{h1} \geq 55.396$ per year. Figure 2(b) identifies two control scenarios involving the treatment rate for recurrent infections ($\phi_{h2}$). When $\beta_{hv} \leq 232.323$, it suffices that $\phi_{h2} \leq 68.832$ to achieve $\mathcal{R}_0 < 1$. However, if $\phi_{h2} \leq 16.840$, the admissible range of $\beta_{hv}$ expands to $\beta_{hv} \leq 539.394$. Figure 2(c) demonstrates that a broad range of treatment completion proportions ($\varepsilon$)

*Corresponding author: Calistus Ngonghala, can ensure control. Specifically, when $\beta_{hv} \leq 818.182$, $\varepsilon \geq 0.561$ is sufficient, whereas for $\beta_{hv} \approx 1496.97$, a much higher completion level ($\varepsilon \geq 0.970$) is required. These findings underscore the dominant role of mosquito biting intensity: beyond sufficiently high $\beta_{hv}$ values, reducing $\mathcal{R}_0$ below unity becomes infeasible. Figure 2(d) reveals a wide admissible region for the progression rates from asymptomatic to symptomatic infection. In particular, when $\psi_{sa} \geq 33.382$, it is necessary that $\psi_{wa} \geq 3.830$ to ensure $\mathcal{R}_0 < 1$. Figure 2(e) highlights the joint impact of $\beta_{hv}$ and antimalarial drug resistance. For $\beta_{hv} \approx 513.131$, resistance must not exceed 0.475, whereas for $\beta_{hv} \leq 920.202$, it must be at most 0.0354. If $\beta_{hv} > 920.202$, control is unattainable regardless of resistance levels, indicating that both parameters must remain sufficiently low. Figure 2(f)–(g) illustrate the potential for malaria elimination through combined interventions using insecticide-treated nets (ITNs) and indoor residual spraying (IRS), consistent with Figure 4(f)–(g) in the main text. Figure 2(h) shows that multiple combinations of human recruitment ($\Lambda_h$) and mosquito mortality ($\mu_v$) yield $\mathcal{R}_0 < 1$. For example, when $\Lambda_h \approx 1.2 \times 10^7$, $\mu_v \geq 20.758$ is required, whereas for $\Lambda_h \approx 3.818 \times 10^7$, $\mu_v \leq 33.182$ suffices. Finally, Figure 2(i) indicates that acceptable combinations of $\mu_v$ and the asymptomatic transmission modification factor ($\theta$) exist. When $\mu_v = 26.707$, $\theta \leq 0.394$, while for $\mu_v \geq 32.576$, $\theta \leq 0.0960$. Thus, increasing $\mu_v$ enlarges the admissible range of $\theta$ consistent with disease control.

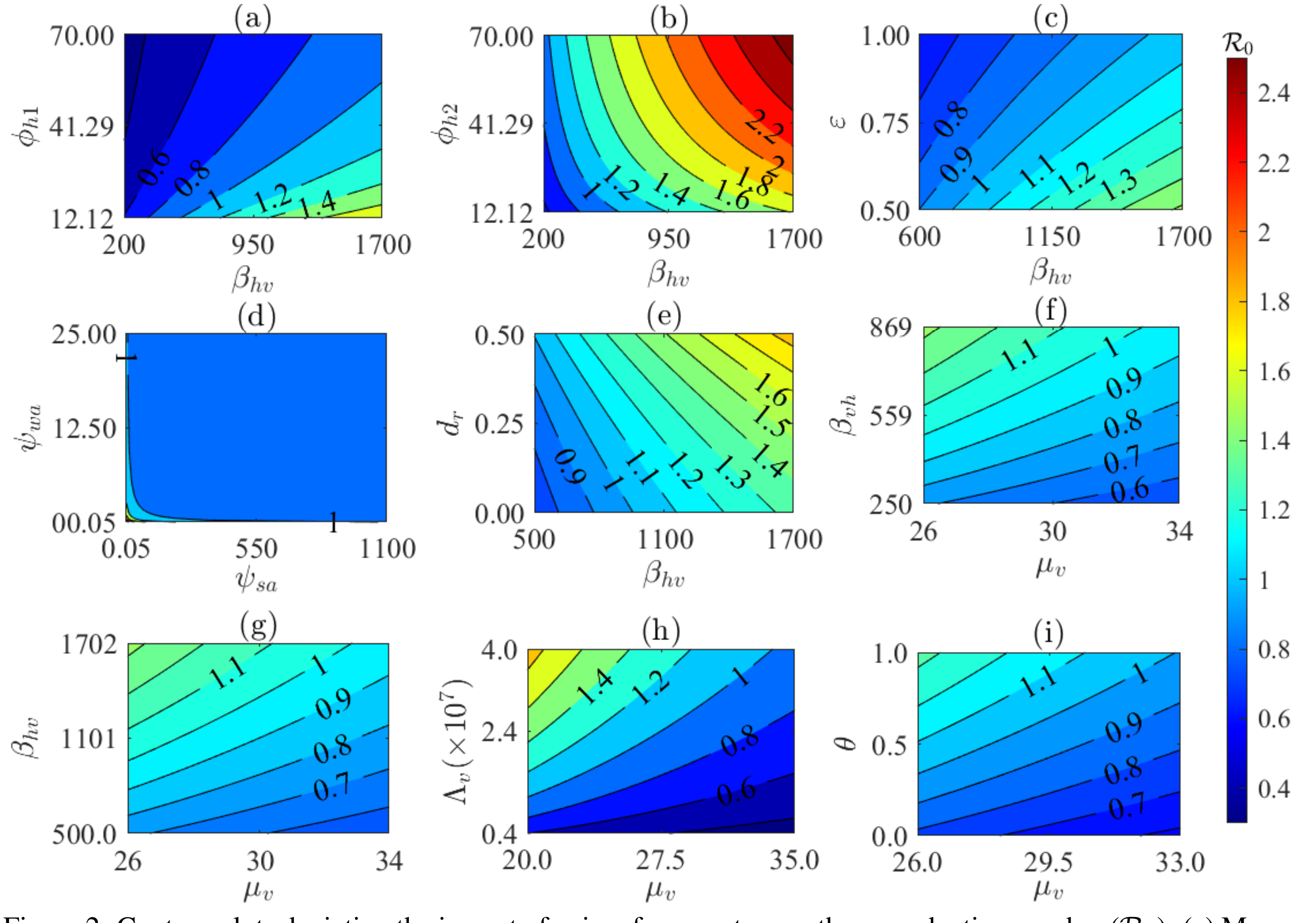


Figure 2: Contour plots depicting the impact of pairs of parameters on the reproduction number ($\mathcal{R}_0$). (a) Mosquito biting rate on infectious humans ($\beta_{hv}$) and malaria treatment rate ($\phi_{h1}$), (b) $\beta_{hv}$ and average duration for individuals without treatment to become asymptomatic ($\phi_{h2}$), (c) $\beta_{hv}$ and proportion of individual who receive optimal treatment ($\varepsilon$), (d) $\psi_{wa}$ and $\psi_{sa}$, (e) $\beta_{hv}$ and drug resistance ($d_r$), (f) $\beta_{vh}$ and $\mu_v$, (g) $\beta_{hv}$ and $\mu_v$, (h) $\Lambda_v$ and $\mu_v$, and (i) $\mu_v$ and $\theta$. Apart from the varied parameters, the other parameters used are those for Nigeria presented in the last columns of Tables 3-4 of the main text.

#### 3. Assessing the impact of (optimum and sub-optimum) treatment on malaria dynamics in Nigeria

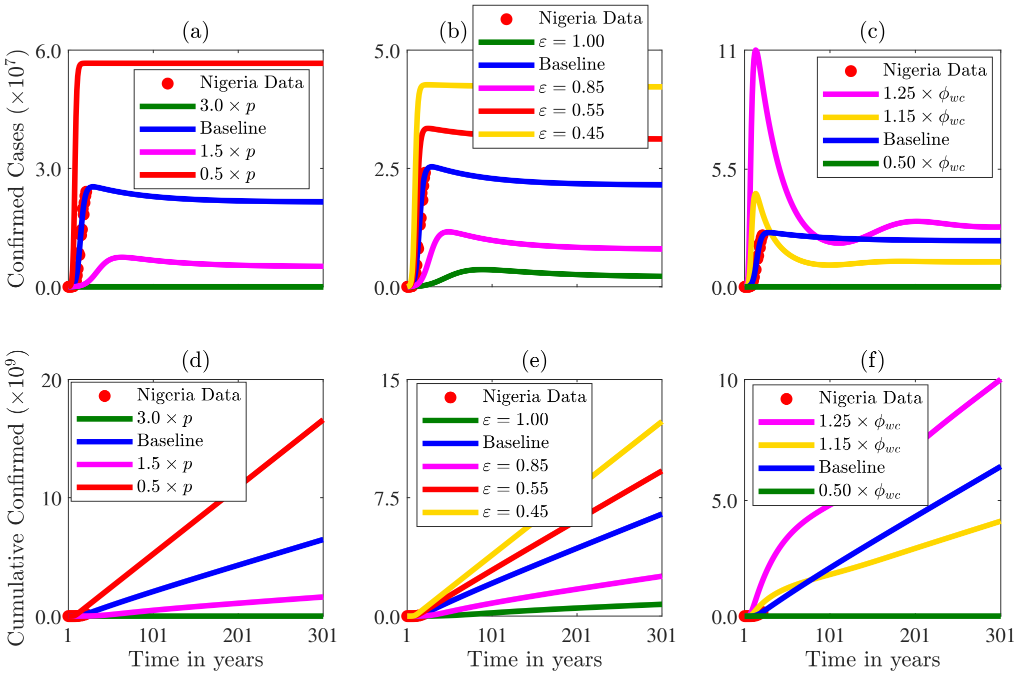


Figure 3: Simulations of the model depicting the impact on the confirmed annual cases of the (a) proportion of individuals who seek treatment ($p$), (b) proportion of individuals who complete optimum treatment ($\varepsilon$), and (c) the sub-optimum treatment rate ($\phi_{wc}$). Corresponding cumulative results are presented in (d)-(f). The other parameter values used for the simulations are those on Nigeria and presented in Tables 3-4.

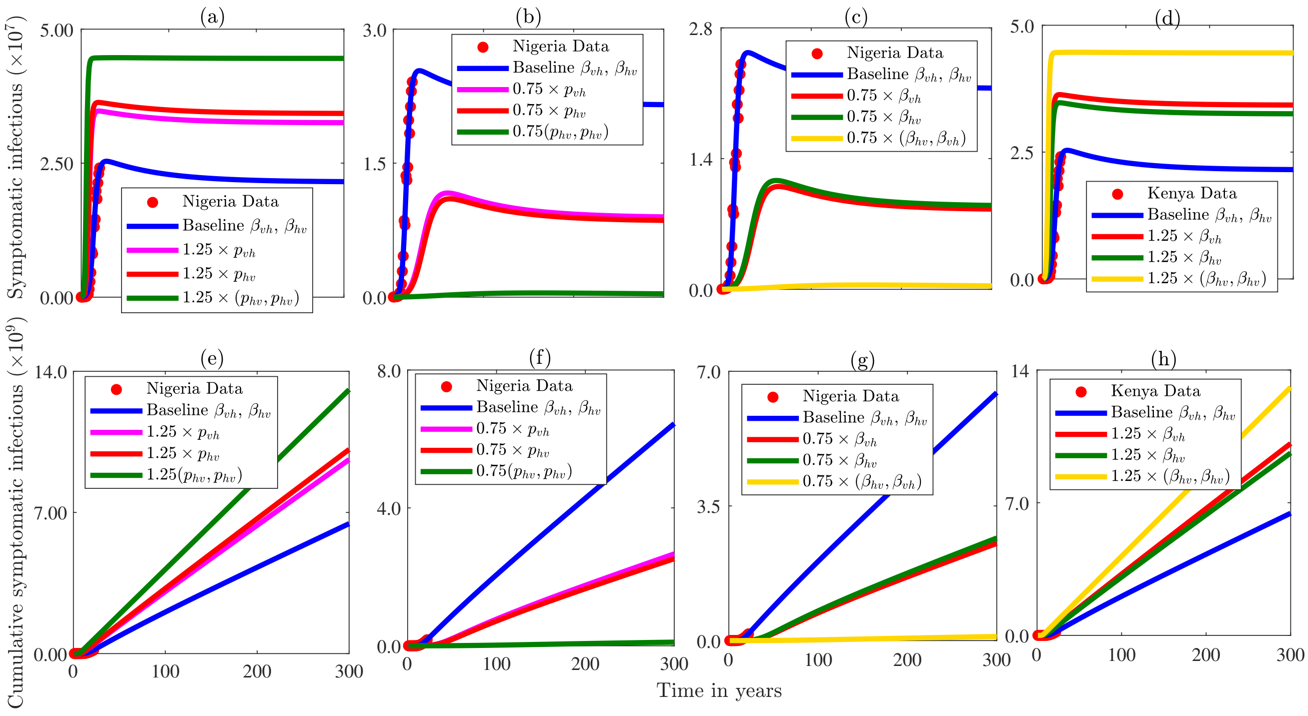

Figure 4: Illustration of the effects of varying the human-to-mosquito transmission probability ($p_{hv}$), the mosquito-to-human transmission probability ($p_{vh}$) (panels (a)–(b)), and the mosquito biting rates $\beta_{vh}$ and $\beta_{hv}$ (panel (c)-(d)) on the annual number of exposed humans. Panels (e)–(h) show the corresponding cumulative exposed populations. All other parameter values used in the simulations are for Kenya and presented in Tables 3 and 4 of the main paper.

#### 4. The initial conditions of the fitting and all time series simulations

The initial conditions are given in the table below.

Table 1: Initial Condition for the state variables per country

| Variables | Kenya | Nigeria |
|---|---|---|
| $S_h$ | $33,420,342$ | $133,469,764$ |
| $E_h$ | $10$ | $50$ |
| $I_h$ | $20,905$ | $400$ |
| $R_w$ | $200,000$ | $50$ |
| $E_w$ | $100$ | $30$ |
| $I_{wc}$ | $7,239$ | $892$ |
| $I_{wa}$ | $10$ | $10$ |
| $I_{sa}$ | $2,000$ | $30$ |
| $R_s$ | $12$ | $20$ |
| $E_s$ | $330$ | $10$ |
| $I_{sc}$ | $1,572$ | $600$ |
| $S_v$ | $6,188,952$ | $31,039,480$ |
| $E_v$ | $200$ | $400$ |
| $I_v$ | $9,000$ | $800$ |

#### 5. Sensitivity and uncertainty analysis of the model parameters on dynamics on Malaria in Nigeria

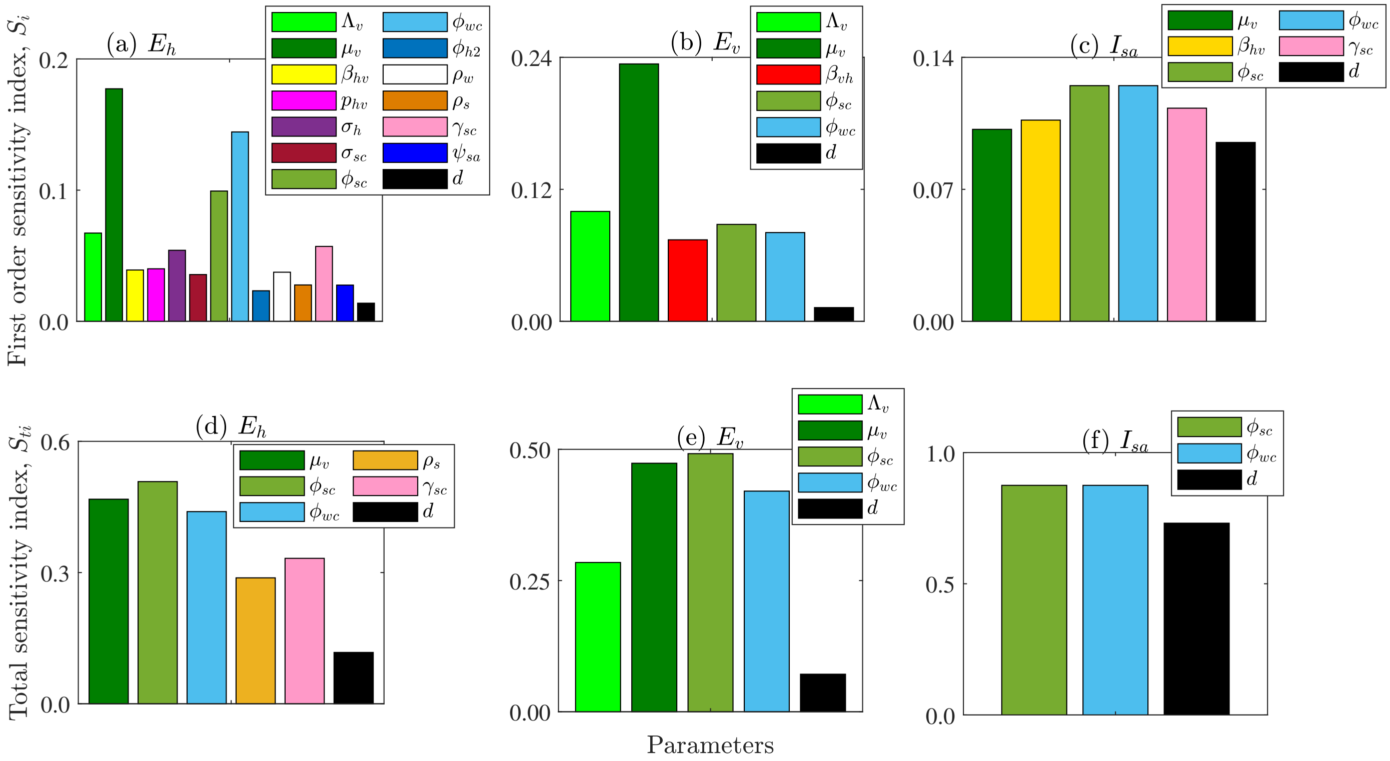


Figure 5: Global sensitivity analysis of the model using the extended Fourier Amplitude Sensitivity Test (eFAST) approach. First order sensitivity of the (a) exposed human population ($E_h$), (b) exposed mosquito population ($E_v$), and (c) asymptomatic infectious malaria cases resulting from sub-optimum treatment ($I_{sa}$), with respect to the parameters of the model. The indices displayed are calculated at time, $t = 40$. Only the sensitivities of parameters that are significantly different from the dummy variable ($d$) with a p-value less than 0.05 are displayed. The initial condition and baseline parameters on Nigeria (in the main paper) were used for this figure.
